# Cardiovascular disease and severe hypoxemia associated with higher rates of non-invasive respiratory support failure in COVID-19

**DOI:** 10.1101/2020.09.27.20202747

**Authors:** Jing Gennie Wang, Bian Liu, Bethany Percha, Stephanie Pan, Neha Goel, Kusum S. Mathews, Cynthia Gao, Pranai Tandon, Max Tomlinson, Edwin Yoo, Daniel Howell, Elliot Eisenberg, Leonard Naymagon, Douglas Tremblay, Krishna Chokshi, Sakshi Dua, Andrew Dunn, Charles Powell, Sonali Bose

**Affiliations:** Division of Pulmonary, Critical Care Medicine and Sleep Medicine, Department of Medicine, Icahn School of Medicine at Mount Sinai, New York, NY, USA; Division of Pulmonary, Critical Care Medicine and Sleep Medicine, Department of Medicine, The Ohio State University Wexner Medical Center, Columbus, OH, USA; Department of Population Health Science and Policy, Icahn School of Medicine at Mount Sinai, New York, NY, USA; Department of Genetics and Genomic Sciences, Icahn School of Medicine at Mount Sinai, New York, NY, USA; Division of General Internal Medicine, Department of Medicine, Icahn School of Medicine at Mount Sinai, New York, NY, USA; Department of Emergency Medicine, Icahn School of Medicine at Mount Sinai, New York, NY, USA; Department of Medicine, Icahn School of Medicine at Mount Sinai, New York, NY, USA; Division of Hematology and Oncology, Department of Medicine, Icahn School of Medicine at Mount Sinai, New York, NY, USA; Division of Hospital Medicine, Department of Medicine, Icahn School of Medicine at Mount Sinai, New York, NY, USA

**Author notes:** Corresponding author: Dr. Sonali Bose, One Gustave L. Levy Place, Box 1232, New York, NY 10029. **Conflicts of interest**: None. JGW, BP, SP, BL and SB take responsibility for the integrity of the data and accuracy of the analyses. SB, JGW and BP conceived the original idea and designed the study. JGW, CG, MT, PT, EY, DH, EE, LN, DT, KC, and SB contributed to the data collection. SP, BL and BP performed the statistical analyses. JGW, BP, SP, BL, NG, KSM, SD, AD, CP, and SB contributed to data interpretation. JGW, BL, and SB drafted the manuscript. All authors revised the manuscript for intellectual content and approved the final version of the manuscript. **Funding source**: None.

**Keywords:** Coronavirus-2019, high-flow oxygen, non-invasive mechanical ventilation, endotracheal intubation, acute hypoxemic respiratory failure, Critical care, respiratory insufficiency, ventilation, coronavirus

## Abstract

**Rationale:** Acute hypoxemic respiratory failure (AHRF) is the major complication of coronavirus disease 2019 (COVID-19), yet optimal respiratory support strategies are uncertain.

**Objectives:** To describe outcomes with high-flow oxygen delivered through nasal cannula (HFNC) and non-invasive positive pressure ventilation (NIPPV) in COVID-19 AHRF and identify individual factors associated with failure.

**Methods:** We performed a retrospective cohort study of hospitalized adults with COVID-19 treated with HFNC and/or NIPPV to describe rates of success (live discharge without endotracheal intubation (ETI)), and identify characteristics associated with failure (ETI and/or in-hospital mortality) using Fine-Gray sub-distribution hazard models.

**Results:** A total of 331 and 747 patients received HFNC and NIPPV as the highest level of non-invasive respiratory support, respectively; 154 (46.5%) in the HFNC cohort and 167 (22.4%) in the NIPPV cohort were successfully discharged without requiring ETI. In adjusted models, significantly increased risk of HFNC and NIPPV failure was seen among patients with cardiovascular disease (subdistribution hazard ratio (sHR) 1.82; 95% confidence interval (CI), 1.17-2.83 and sHR 1.40; 95% CI 1.06-1.84), respectively, and among those with lower oxygen saturation to fraction of inspired oxygen (SpO_2_/FiO_2_) ratio at HFNC and NIPPV initiation (sHR, 0.32; 95% CI 0.19-0.54, and sHR 0.34; 95% CI 0.21-0.55, respectively).

**Conclusions:** A significant proportion of patients receiving non-invasive respiratory modalities for COVID-19 AHRF achieved successful discharge without requiring ETI, with lower success rates among those with cardiovascular disease or more severe hypoxemia. The role of non-invasive respiratory modalities in COVID-19 related AHRF requires further consideration.

## INTRODUCTION

The severe acute respiratory syndrome coronavirus 2 (SARS-CoV-2) is responsible for the coronavirus disease 2019 (COVID-19) pandemic, resulting in over 16.5 million cases and more than 655,000 deaths across 216 countries (1). The burden of cases in the United States has exceeded that of any other country, with New York City as the epicenter of the pandemic at the time of this study, accounting for over 226,000 cases as of July 30, 2020 (2). The hallmark of COVID-19 is the development of acute respiratory illness, with a significant number of hospitalized adult patients progressing to acute respiratory distress syndrome (ARDS) (3-6), and up to 18% ultimately requiring endotracheal intubation (ETI) and invasive mechanical ventilation (IMV) (5-9). Importantly, mortality rates after ETI are substantial, ranging from 65-96% (3, 4, 7, 10).

While some experts during the pandemic have advocated for early ETI and invasive mechanical ventilation (IMV) (11), the optimal respiratory support strategy for COVID-19 acute hypoxemic respiratory failure (AHRF) remains unclear (12). Compared to low-flow supplemental oxygen, HFNC improves oxygenation and work of breathing (13) and reduces ETI rates (14-16), while NIPPV may be associated with a lower risk of both ETI and mortality in certain non-COVID-19 populations with AHRF (16). Despite these benefits, some studies have shown that treatment failure with HFNC and NIPPV is associated with worse outcomes, including increased mortality, possibly related to delayed ETI and initiation of IMV (17-19). Reports of outcomes with HFNC and NIPPV in the COVID-19 population, however, remain limited to small case series (20, 21), with significant variability in use of these modalities across centers (3, 9, 10, 22). Accordingly, individual characteristics associated with HFNC and NIPPV failure in COVID-19 AHRF are unknown. Unclear benefits of HFNC and NIPPV have resulted in some experts advocating for avoidance of these modalities in favor of early ETI during the pandemic (23).

To address this knowledge gap, we sought to describe patterns of HFNC and NIPPV use and outcomes for patients admitted with COVID-19 AHRF within a large New York City academic health system. We subsequently identified specific demographic and clinical characteristics at the time of HFNC and NIPPV initiation that were associated with HFNC and NIPPV failure, highlighting patient subgroups particularly susceptible to poor outcomes with non-invasive respiratory support.

## METHODS

### Study design and participants

This retrospective cohort study included patients admitted to one of five hospitals within the Mount Sinai Health System, New York City, including a large quaternary referral center, a tertiary care hospital, and three community hospitals, totaling 2,590 beds. The Icahn School of Medicine at Mount Sinai institutional review board approved the study, and the requirement for informed consent was waived due to minimal risk.

For the descriptive study aim, we included all hospitalized patients between >18 and 100 years of admitted between March 1 and April 28, 2020 with a positive SARS-CoV-2 reverse transcriptase polymerase chain reaction of nasopharyngeal swab samples, and who required HFNC and/or NIPPV prior to achieving an outcome of first ETI, live hospital discharge, or in-hospital mortality (Figure 1). Patients were followed up to May 31, 2020. Patients from the descriptive population without missing covariates were included in the analytic subset (Figure 1). For transferred patients, defined as discharged from one of five hospital sites with readmission at another within 12 hours, both admissions were included as a single hospitalization. If a patient was discharged for a period of >12 consecutive hours and subsequently readmitted, only the first admission was considered.

**Figure 1.**
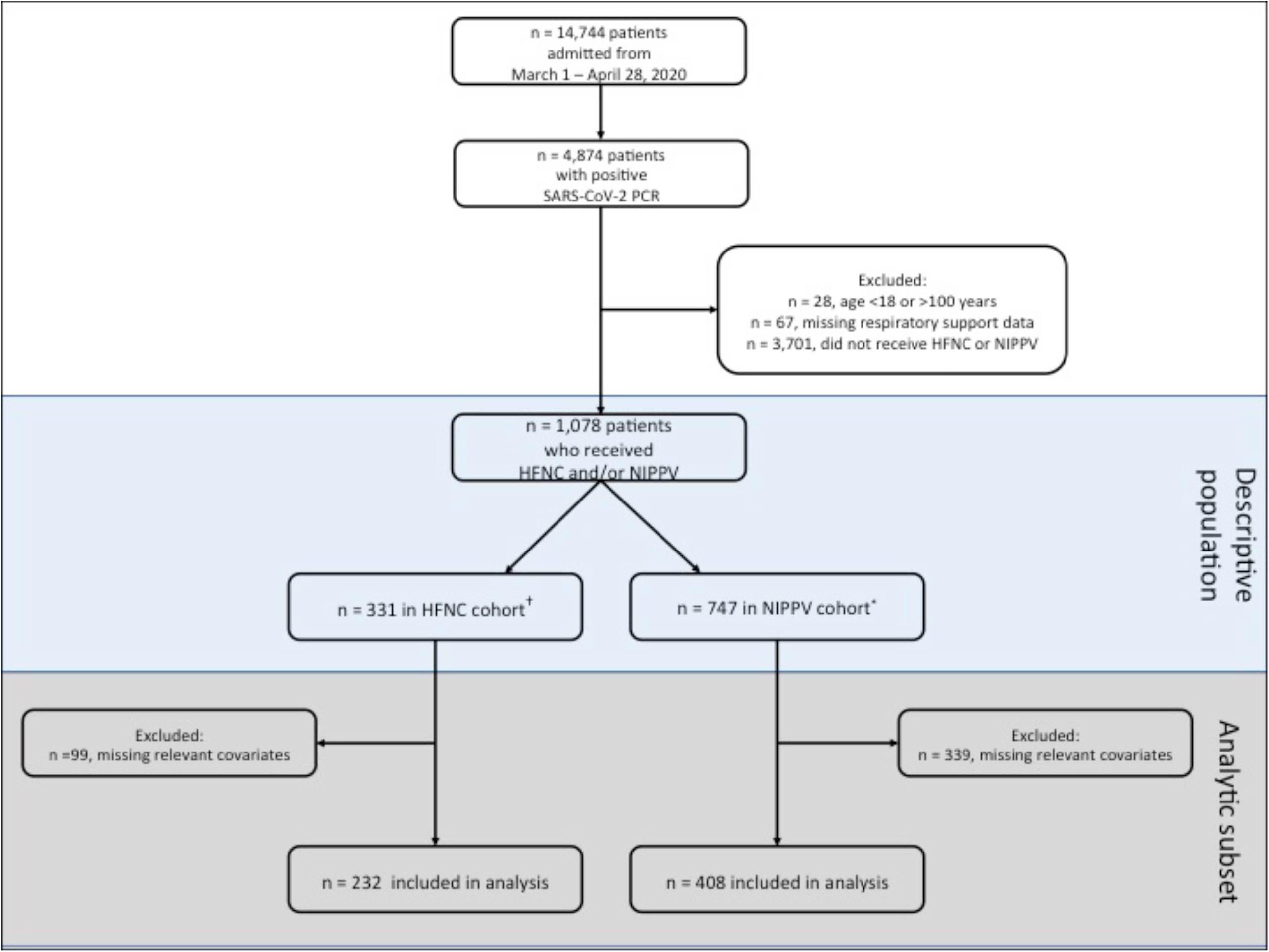
Selection flow diagram for study participant inclusion. ^*^Includes patients who received HFNC as the highest non-invasive respiratory support prior to outcome of first endotracheal intubation, live hospital discharge or in-hospital mortality. ^†^Includes patients who received NIPPV as the highest non-invasive respiratory support prior to outcome of first endotracheal intubation, live hospital discharge or in-hospital mortality. SARS-CoV-2 PCR: Severe acute respiratory coronavirus-2 polymerase chain reaction; HFNC: High-flow oxygen delivered through nasal cannula; NIPPV: Non-invasive positive pressure ventilation.

Data were extracted from electronic databases, including Epic Clarity and Epic Caboodle (Epic Systems Corporation). ETI timings for 400 patients were validated using chart review; only 3 out of 400 (0.75%) showed inconsistencies with the automatically extracted data due to charting errors. Data collected included hospital disposition (either live discharge or death), patient demographics, code status of do-not-intubate (DNI), comorbidities (captured using ICD-10 codes), medications (including anticoagulation therapies and corticosteroids), vital signs, laboratory tests, and treatment with respiratory support devices (nasal cannula, face mask, non-rebreather mask, HFNC, NIPPV (defined as continuous or bilevel positive pressure devices), and/or ETI with IMV). The last recorded vital signs and laboratory values prior to device initiation were selected; if these were not available (for example, if the patient was placed on a device immediately at the time of admission) the measurements occurring soonest after device initiation were selected.

### Outcomes

The primary outcome was treatment success, defined as live hospital discharge without need for ETI. Treatment failure was defined as ETI and/or in-hospital mortality, whichever occurred first, to account for patients who may have died before ETI documentation (e.g. during cardiopulmonary resuscitation), or for whom IMV may have been indicated but who died without ETI due to DNI code status. Secondarily, in multivariable models including the subset of patients with non-missing data for covariates included in the main model, we explored specific demographic and clinical characteristics at the time of HFNC or NIPPV initiation that were associated with treatment failure.

### Non-invasive respiratory support cohorts

To simplify multiple transitions between HFNC and NIPPV that often take place during hospitalization, each patient was categorized into one of two cohorts based on the highest level of non-invasive respiratory support required prior to outcome (as defined above), or if no outcome was observed, the end of the study period. Accordingly, the HFNC cohort included patients who received HFNC but never NIPPV, while the NIPPV cohort included patients who received NIPPV, or both HFNC and NIPPV at any point between hospital admission and outcome, irrespective of sequence. In addition, timing of initiation and total duration of HFNC and/or NIPPV use from hospital admission to an outcome were calculated.

### Covariates

Patient demographics included age, sex, race/ethnicity, and insurance type, as well as hospital site. Clinical factors included comorbidities, smoking status, body mass index (BMI), and DNI code status (if documented prior to HFNC or NIPPV initiation), as well as vital signs, ratio of peripheral blood oxygen saturation to fraction of inspired oxygen (SpO_2_/FiO_2_), and laboratory tests at the time of HFNC or NIPPV initiation. Enrollment into investigational drug trials for COVID-19 was not included as a covariate, as only a portion of those enrolled ultimately received the investigational agent and data regarding specific therapies could not be reported due to embargo.

### Statistical Analyses

Baseline demographics and clinical characteristics at the time of device initiation are presented for both HFNC and NIPPV cohorts. Frequencies and proportions are described for categorical variables and median and interquartile range (IQR) used for continuous measures. We determined detailed follow-up time by outcome subgroups. For each cohort, Fine-Gray competing risk regression models were performed at univariable and multivariable levels to estimate the sub-distribution hazard ratios (sHR) with corresponding 95% confidence intervals (CI) of treatment failure, treating live hospital discharge as a competing risk. Patients were censored if they did not experience an outcome by end of the study period. Covariates in the univariable models with p ≤0.15 were entered into multivariable models for each cohort (Table E1). All continuous variables used in models, except for age and temperature, were natural-log transformed to address observed skew in their underlying distributions (a standard procedure in regression models).

We performed the following sensitivity analyses to test the robustness of the outcomes from the main model: (1) Addition of prophylactic- and therapeutic-dose anticoagulation and/or corticosteroids administered prior to device initiation as individual covariates in the multivariable models, as these interventions may have impacted the risk of failure and clinical characteristics may also differ by treatment status; (2) excluding interleukin-6 and BMI, the two covariates with the most missing values, from the multivariable models to increase sample size and better represent the initial study population; and (3) evaluating differences in demographics and clinical characteristics between patients with and without any missing covariates (irrespective of whether they were included in adjusted models). P-values <0.05 were considered statistically significant. Analyses were performed using SAS version 9.4 (SAS Institute Inc., Cary, NC), and R (Version 3.5.0) using Rstudio (Version 1.1.453, RStudio, Inc.).

## RESULTS

### Population characteristics

Of the 1,078 total patients receiving high-level non-invasive respiratory support at any point during their hospitalization, 331 (30.7%) and 747 (69.3%) required HFNC and NIPPV, respectively, as their highest level of non-invasive respiratory support with almost half of the NIPPV group receiving both HFNC and NIPPV (316 patients, 42.3%) (Table 1). There was significant variability in HFNC and NIPPV use across hospital sites, ranging from 3.5 to 9.5% for HFNC and 11.0 to 21.6% for NIPPV.

**Table 1.** Patient demographics, characteristics and treatments received at the time of HFNC and NIPPV initiation

|  | HFNC cohort<br>(n = 331) | NIPPV cohort<br>(n = 747) |
| --- | --- | --- |
| **Age (years), median (IQR)** | 67.7 (57.8, 78) | 69.4 (60.9, 79.5) |
| **Male sex, no. (%)** | 200 (60.4) | 466 (62.4) |
| **Race, no. (%)** |  |  |
| White | 78 (23.6) | 174 (23.3) |
| Black | 84 (25.4) | 185 (24.8) |
| Asian/Pacific Islander | 19 (5.7) | 40 (5.4) |
| Other/unknown | 150 (45.3) | 348 (46.6) |
| **Ethnicity, no. (%)** |  |  |
| Non-Hispanic | 193 (58.3) | 442 (59.2) |
| Hispanic | 101 (30.5) | 216 (28.9) |
| Unknown | 37 (11.2) | 89 (11.9) |
| **Insurance, no. (%)** |  |  |
| Medicare | 134 (40.5) | 315 (42.2) |
| Medicaid | 48 (14.5) | 89 (11.9) |
| Private | 66 (19.9) | 136 (18.2) |
| Self-pay | 5 (1.5) | 24 (3.2) |
| Other/unknown | 78 (23.6) | 183 (24.5) |
| **Comorbidities, no. (%)** |  |  |
| Hypertension | 167 (50.5) | 406 (54.4) |
| Type 2 diabetes mellitus | 112 (33.8) | 287 (38.4) |
| Chronic lung disease* | 192 (58) | 379 (50.7) |
| Cardiovascular disease† | 168 (50.8) | 407 (54.5) |
| Cerebrovascular disease | 38 (11.5) | 73 (9.8) |
| Liver disease | 17 (5.1) | 52 (7) |
| Chronic kidney disease | 52 (15.7) | 149 (20) |
| Cancer | 35 (10.6) | 84 (11.2) |
| **Smoking status, no. (%)** |  |  |
| Never | 168 (50.8) | 360 (48.2) |
| Unknown | 70 (21.2) | 172 (23) |
| Former | 81 (24.5) | 187 (25) |
| Current | 12 (3.6) | 28 (3.8) |
| **Vital signs, median (IQR)** |  |  |
| Heart rate (beats per minute) | 94 (82, 106) | 95 (80, 109) |
| Systolic blood pressure | 125 (113, 143) | 128 (115, 144) |
| Diastolic blood pressure | 73 (64, 80) | 73 (64, 81) |
| Respiratory rate | 20 (19, 25) | 22 (20, 28) |
| SpO<sub>2</sub>/FiO<sub>2</sub> ratio | 98 (93, 125) | 97 (92, 121.4) |
| Temperature (°Fahrenheit) | 98.4 (97.5, 99.5) | 98.4 (97.6, 99.3) |
| **Body mass index, median (IQR)** | 28.8 (25.1, 34.0) | 28.7 (25.4, 34.3) |
| **Laboratory values, median (IQR)** |  |  |
| White blood cell count (x10<sup>3</sup>/μL) | 9.0 (6.7, 11.9) | 9.8 (7.2, 13.4) |
| Peripheral lymphocyte (%) | 9.6 (6.1, 14.3) | 8.7 (5.6, 13.2) |
| Hemoglobin (g/dL) | 12.7 (11.2, 14) | 12.9 (11.2, 14.3) |
| Platelets (x10<sup>3</sup>/μL) | 249 (175, 330) | 236 (171, 300) |
| D-Dimer (μg/mL) | 1.8 (1.0, 5.1) | 2.5 (1.3, 5.8) |
| C-reactive protein (mg/L) | 170.8 (94.9, 239.4) | 187.1 (102.7, 265.4) |
| Ferritin (ng/mL) | 1090.0 (597.5, 2485.0) | 1132.0 (509.0, 2276.0) |
| Interleukin-6 (pg/mL) | 83.7 (40.6, 171.5) | 93.0 (45.5, 207.0) |
| Pro-calcitonin (ng/mL) | 0.3 (0.1, 0.8) | 0.4 (0.2, 1.2) |
| Creatinine (mg/dL) | 0.9 (0.7, 1.6) | 1.2 (0.8, 1.9) |
| **Code status<sup>†</sup>** |  |  |
| Do-not-intubate, no. (%) | 59 (17.8) | 96 (12.9) |
| **Treatments received<sup>§</sup>** |  |  |
| Prophylactic anticoagulation, no. (%) | 119 (36) | 352 (47.1) |
| Therapeutic anticoagulation, no. (%) | 95 (28.7) | 193 (25.8) |
| Corticosteroids, no. (%) | 5 (1.5) | 12 (1.6) |

| Hospital admission site, no. (%) |  |  |
| --- | --- | --- |
| Quaternary referral hospital | 152 (45.9) | 190 (25.4) |
| Community hospital A | 31 (9.4) | 199 (26.6) |
| Community hospital B | 28 (8.5) | 95 (12.7) |
| Community hospital C | 78 (23.6) | 177 (23.7) |
| Tertiary hospital | 42 (12.7) | 86 (11.5) |

\*Chronic lung disease includes: Asthma, emphysema, chronic obstructive pulmonary disease, chronic bronchitis, bronchiectasis, atelectasis, diaphragmatic disease, interstitial lung diseases, sarcoidosis, and disorders of mediastinum.
†Cardiovascular disease includes: Atherosclerotic heart disease, ischemic heart disease, congestive heart failure, rheumatic heart disease, pericardial disease, myocarditis, endocarditis, valvular disorders, cardiomyopathy, arrhythmias, history of cardiac arrest, peripheral vascular disease, aortic aneurysm, orthostatic hypotension, pulmonary hypertension, cardiac arrest, post-procedural cardiac complications.
‡Status at time of initiation of HFNC or NIPPV
§From hospital admission to initiation of HFNC or NIPPV.
Missing sample size in the HFNC cohort included 38 for SpO<sub>2</sub>/FiO<sub>2</sub> ratio, 41 for body mass index, 3 for peripheral lymphocyte percent, 16 for D-Dimer, 13 for C-reactive protein, 11 for ferritin, 72 for interleukin-6, and 21 for pro-calcitonin.
Missing sample size in the NIPPV cohort included 52 for SpO<sub>2</sub>/FiO<sub>2</sub> ratio, 132 for body mass index, 5 for peripheral lymphocyte percent, 40 for D-Dimer, 35 for C-reactive protein, 37 for ferritin, 232 for interleukin-6, and 82 for pro-calcitonin.
HFNC: High-flow oxygen delivered through nasal cannula; NIPPV: Non-invasive positive pressure ventilation; IQR: Interquartile; SpO<sub>2</sub>: Oxygen saturation; FiO<sub>2</sub>: Fraction of inspired oxygen.

Demographic and clinical characteristics of patients in the HFNC and NIPPV cohorts are presented in Table 1. In both cohorts, comorbidities, including chronic lung and cardiovascular diseases (Table E2), were common, and distributions of age, sex, and race/ethnicity were similar. At time of HFNC and NIPPV initiation, patients had a median (IQR) SpO_2_/FiO_2_ of 98 (93, 125) and 97 (92, 121.4), respectively, suggesting severe hypoxemia. Inflammatory markers were elevated in both groups (Table 1).

Considering dynamic respiratory support requirements including room air and low flow supplemental oxygen over time for a given patient, the highest time-varying probability of receiving HFNC was 36% among those in the HFNC cohort, occurring between days 5-6 of admission (Figure 2). Among patients in the NIPPV cohort, the highest time-varying probability of being on NIPPV (31%) occurred within the first day of admission, while HFNC use in the same group was 12% on day 5 of admission (Figure 2).

**Figure 2.**
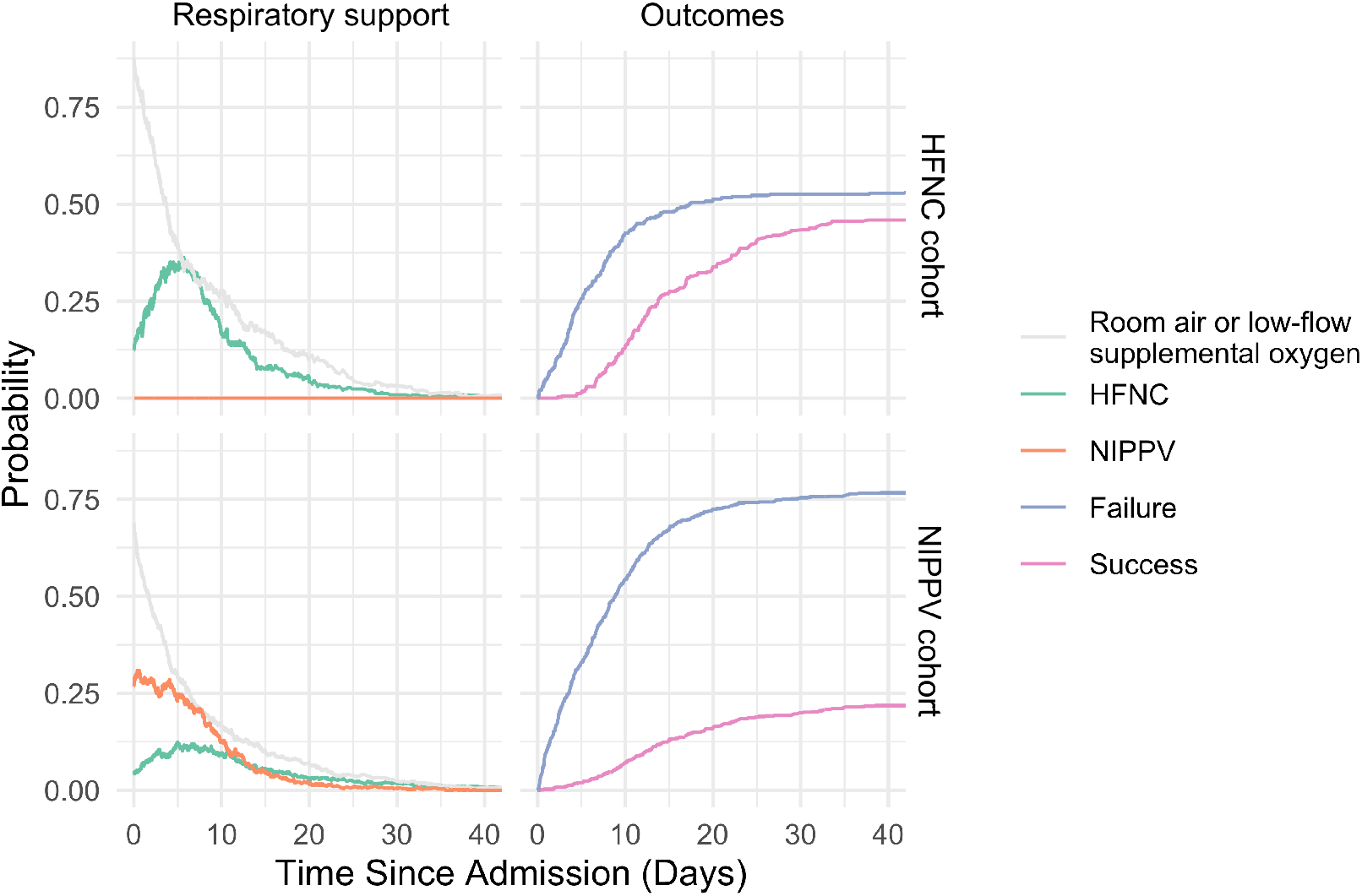
Time-varying probabilities of HFNC and NIPPV use during hospital admission, and corresponding outcome trajectories. Among patients in the HFNC cohort, highest probability of receiving HFNC over the duration of hospital stay was 36%, which occurred between days 5 and 6 of admission, and probabilities of treatment success (live hospital discharge without requiring endotracheal intubation (ETI)), and failure (required ETI and/or in-hospital mortality) were 47% and 54%, respectively. Among patients in the NIPPV cohort, highest probability of receiving NIPPV over the duration of hospital stay was 31%, which occurred within the first day of admission, while the highest probability of receiving HFNC was 12% on day 5 of admission. Probabilities of treatment success and failure for patients in the NIPPV cohort were 22% and 77%, respectively. HFNC: High-flow oxygen delivered through nasal cannula; NIPPV: Non-invasive positive pressure ventilation.

### Outcomes in the HFNC cohort

Out of 331 patients in the HFNC cohort, 154 (46.5%) were successfully discharged without needing ETI during their hospital stay (Figure 3A). Of the 177 (53.5%) who experienced HFNC failure, 100 (56.5%) required ETI and 77 (43.5%) died without ETI. Among the 100 patients who ultimately required ETI, 35 (35.0%) were eventually discharged, 58 (58.0%) died and 7 (7.0%) were censored (Figure 3A). The median (IQR) follow-up time in the overall HFNC cohort from time to device initiation to any of the defined outcomes (failure, success, or the end of study for those who did not experienced failure or successful discharge) was 5.40 (1.00, 10.7) days, and specifically, was 1.36 (0.54, 3.77) days and 10.2 (6.72, 15.5) days among those who experienced treatment failure and successful discharge, respectively.

**Figure 3.**
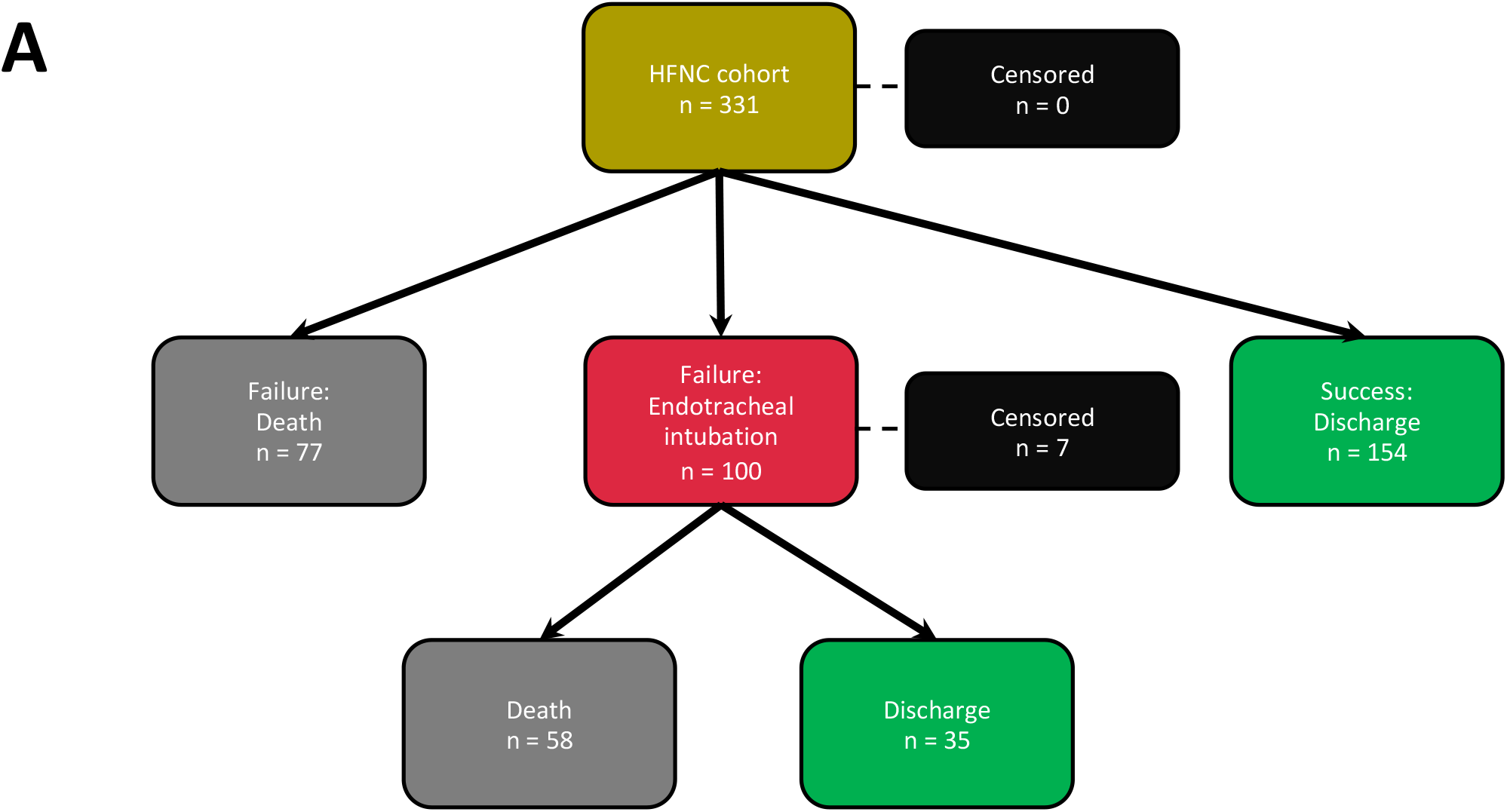
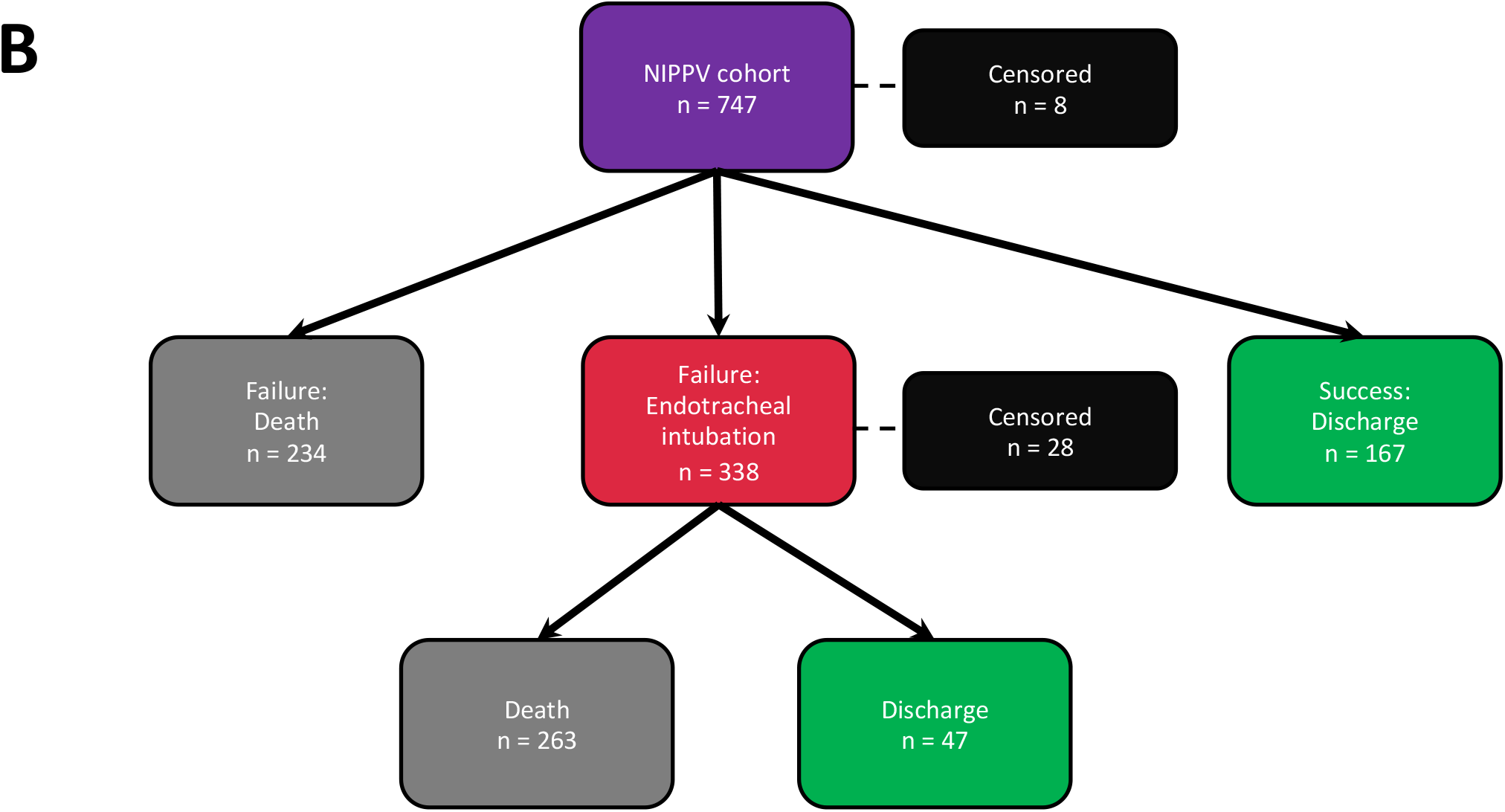
Trajectory of outcomes among patients who were treated with A) HFNC, and B) NIPPV as the highest non-invasive respiratory support at any point during hospitalization prior to outcomes of first endotracheal intubation, live hospital discharge, or in-hospital mortality. Treatment success was defined as live hospital discharge without requiring endotracheal intubation. Treatment failure was defined as requiring endotracheal intubation and/or in-hospital mortality. Patients who did not experience an outcome by the end of the study period were censored. HFNC: High-flow oxygen delivered through nasal cannula; NIPPV: Non-invasive positive pressure ventilation.

### Factors associated with HFNC failure

In multivariable analysis, limited to the subset of patients in the HFNC cohort with complete data for all adjusted covariates (n = 232, 70%), a significantly increased incidence of HFNC failure was observed among patients with cardiovascular disease (sHR 1.82; 95% CI, 1.17-2.83) (Figure 4). A lower SpO_2_/FiO_2_ ratio also increased the risk of HFNC failure (sHR 0.32; 95% CI, 0.19-0.54), as did a lower platelet count (sHR 0.61; 95% CI, 0.43-0.87). Admission to Community hospital C and the tertiary hospital reduced the risk of failure compared to the quaternary referral hospital (sHR 0.55; 95% CI 0.31-0.99, and sHR 0.29; 95% CI, 0.16-0.55, respectively) (Figure 4).

**Figure 4.**
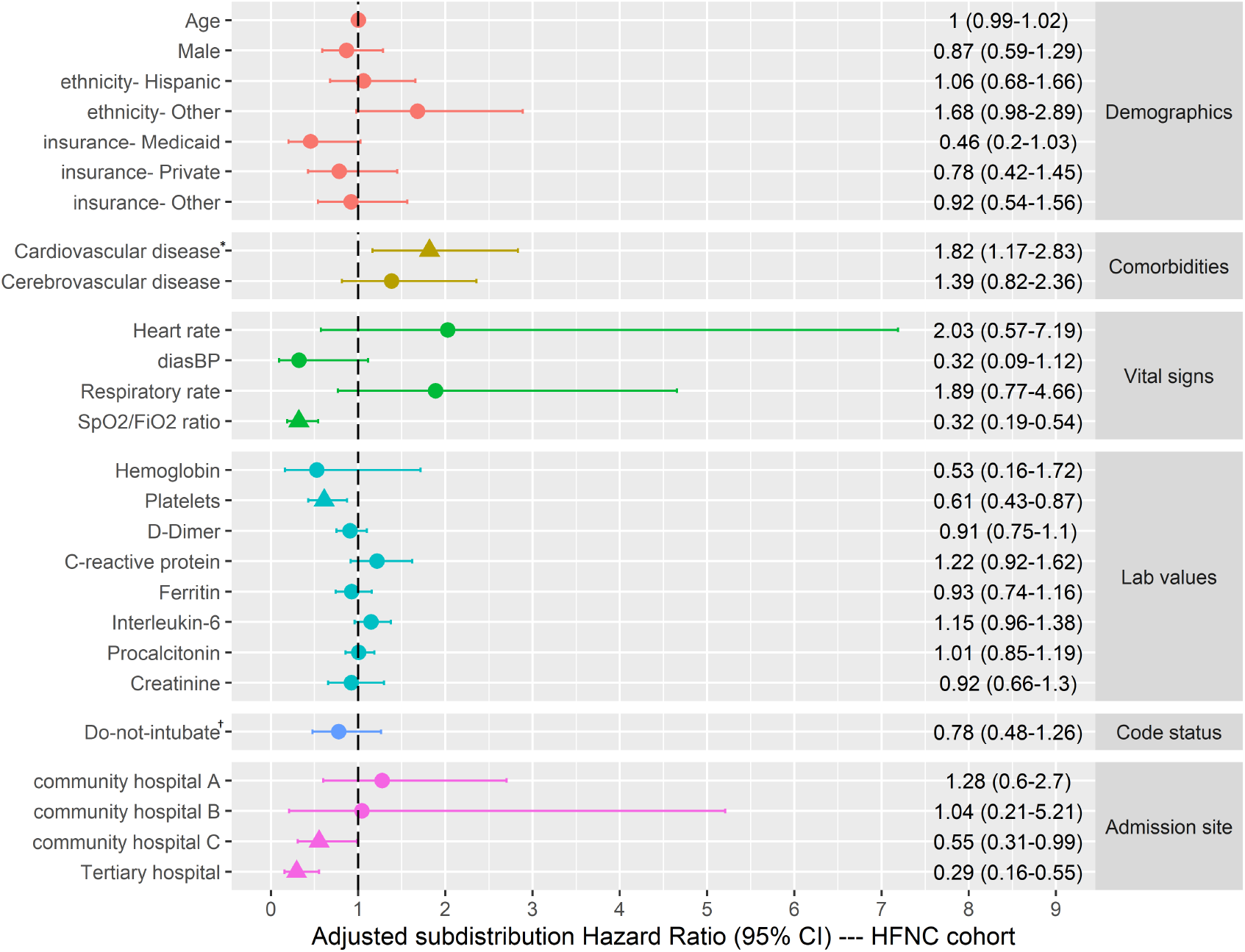
Multivariable analyses with sub-distribution hazard ratio estimates for HFNC failure among the subset of patients in the HFNC cohort with complete covariates included in the multivariable model (n = 232), with live hospital discharge as a competing risk. Statistically significant sHRs are shown with triangles. ^*^Cardiovascular disease includes: Atherosclerotic heart disease, ischemic heart disease, congestive heart failure, rheumatic heart disease, pericardial disease, myocarditis, endocarditis, valvular disorders, cardiomyopathy, arrhythmias, history of cardiac arrest, peripheral vascular disease, aortic aneurysm, orthostatic hypotension, pulmonary hypertension, cardiac arrest, post-procedural cardiac complications. ^†^At time of HFNC initiation. sHR: Sub-distribution hazard ratio; CI: Confidence interval; HFNC: High-flow oxygen through nasal cannula; CI: Confidence interval; SpO_2:_ Oxygen saturation; FiO_2_: Fraction of inspired oxygen. All continuous variables, except for age, were natural log-transformed.

### Outcomes in the NIPPV cohort

Of 747 total patients requiring NIPPV, 167 (22.4%) were successfully discharged without requiring ETI. Of the 572 (76.6%) patients who experienced NIPPV failure, 338 (59.1%) required ETI and 234 (40.9%) died without ETI. Of 338 patients who required ETI, 47 (13.9%) were eventually discharged and 263 (77.8%) died (Figure 3B). The median (IQR) follow-up time for the NIPPV cohort was 4.1 (1.2, 9.1) days, and specifically, was 2.6 (0.8, 6.7) days and 11.2 (6.8, 17.6) days for those with treatment failure and success, respectively.

### Factors associated with NIPPV failure

In multivariable analysis limited to the subset of patients in the NIPPV cohort with complete data for all adjusted covariates (n = 408), co-morbid cardiovascular disease and higher interleukin-6 were associated with increased risk of failure (sHR 1.40; 95% CI, 1.07-1.84, and sHR 1.13; 95% CI 1.02-1.26, respectively) (Figure 5). Increased risk of failure was also associated with a lower SpO_2_/FiO_2_ ratio (sHR 0.34; 95% CI, 0.21-0.55), lower hemoglobin (sHR 0.41; 95% CI, 0.19-0.88), and lower peripheral lymphocyte percentage (sHR 0.74; 95% CI 0.59-0.94).

**Figure 5.**
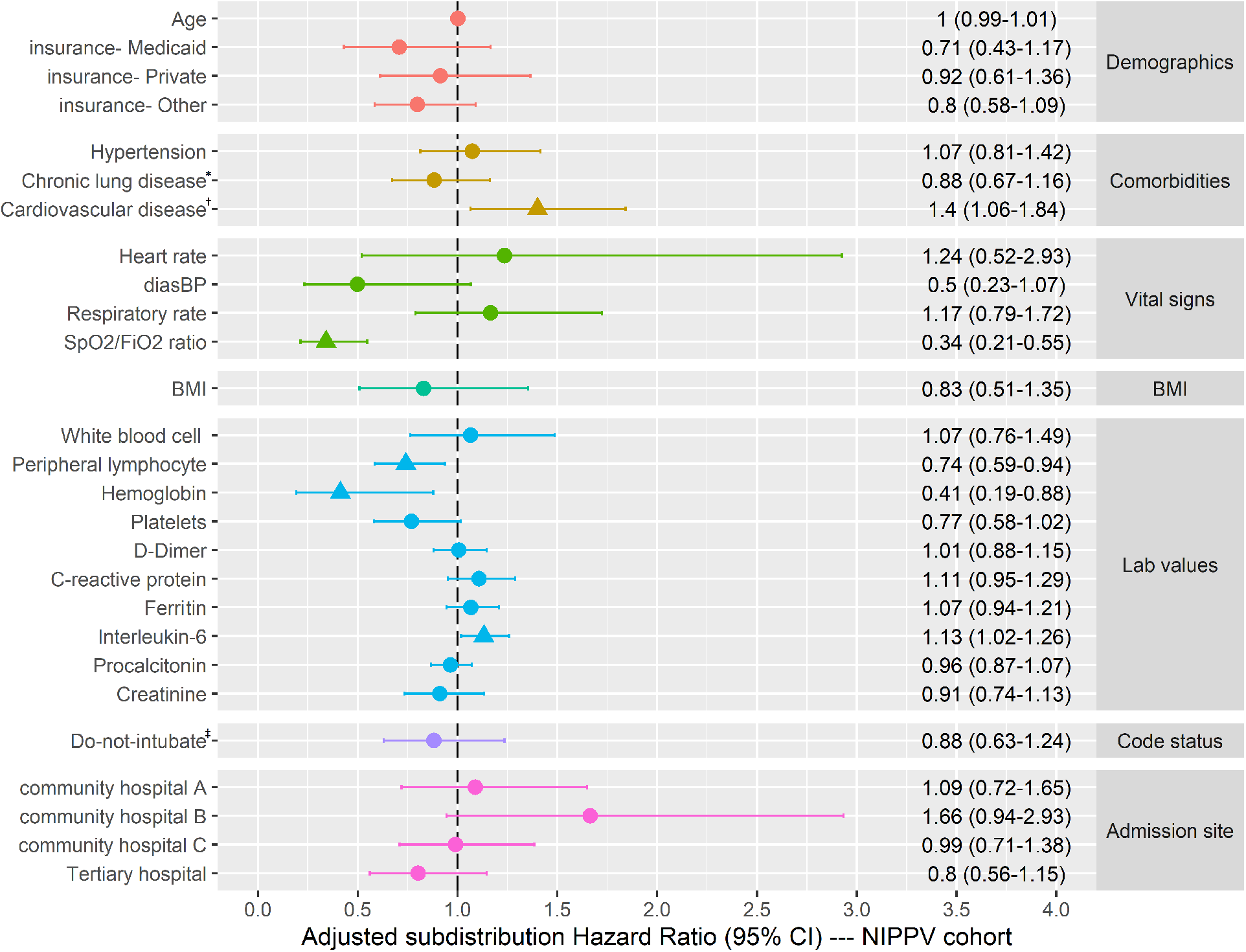
Multivariable analyses with sub-distribution hazard ratio estimates for treatment failure among the subset of patients in the NIPPV cohort with complete covariates included in the multivariable model (n = 408), with live hospital discharge as a competing risk. Statistically significant sHRs are shown in triangles. ^*^Chronic lung disease includes: Asthma, emphysema, chronic obstructive pulmonary disease, chronic bronchitis, bronchiectasis, atelectasis, diaphragmatic disease, interstitial lung diseases, sarcoidosis, and disorders of mediastinum. ^†^Cardiovascular disease includes: Atherosclerotic heart disease, ischemic heart disease, congestive heart failure, rheumatic heart disease, pericardial disease, myocarditis, endocarditis, valvular disorders, cardiomyopathy, arrhythmias, history of cardiac arrest, peripheral vascular disease, aortic aneurysm, orthostatic hypotension, pulmonary hypertension, cardiac arrest, post-procedural cardiac complications. ^‡^At time of NIPPV initiation. sHR: Sub-distribution hazard ratio; CI: Confidence interval; NIPPV: Non-invasive positive pressure ventilation; CI: Confidence interval; SpO_2:_ Oxygen saturation; FiO_2_: Fraction of inspired oxygen. All continuous variables, except for age, were natural log-transformed.

### Sensitivity analyses

In sensitivity analyses where receipt of prophylactic or therapeutic anticoagulation and/or corticosteroids prior to device initiation were added as covariates, co-morbid cardiovascular disease and lower SpO_2_/FiO_2_ ratio remained significantly associated with increased risk of failure in both HFNC (Table E3) and NIPPV cohorts (Table E4). Similarly, despite differences in demographic and clinical characteristics between patients with overall complete vs. incomplete covariate data (Table E5), significant associations between cardiovascular disease and SpO_2_/FiO_2_ ratio remained in models where interleukin-6 and BMI were excluded to encompass a larger subset of patients (Tables E3 and E4).

## DISCUSSION

This is the largest cohort study in patients with COVID-19 AHRF to date to describe patterns of use and outcomes with non-invasive respiratory support modalities of HFNC and NIPPV. We observed that nearly half of the patients treated with HFNC and one-fifth of the patients treated with NIPPV were successfully discharged without requiring ETI. As outcomes with HFNC and NIPPV were variable, we subsequently identified specific clinical features, e.g. co-morbid cardiovascular disease, associated with increased risk of treatment failure (need for ETI and/or in-hospital mortality). Recognition of these characteristics may help to identify vulnerable patient subgroups at higher risk of failure with non-invasive modalities, thus necessitating closer monitoring. Similarly, more severe hypoxemia was associated with a higher rate of treatment failure in those receiving non-invasive strategies, supporting their consideration primarily in patients with milder disease. Dedicated studies to evaluate the efficacy of non-invasive modalities to reduce risks of ETI and mortality in COVID-19 AHRF are clearly warranted.

Limited studies of non-invasive respiratory support outcomes in COVID-19 patients report survival rates of 19.5% to 100% with HFNC (4, 20, 21) and 7.7% to 37.7% with NIPPV (3, 4). Although our aim was not to directly compare these non-invasive modalities, we found that a lower proportion of patients receiving NIPPV were successfully discharged compared to HFNC. A higher failure rate among patients receiving NIPPV may be due to higher illness severity or inherent differences in physiologic mechanisms of HFNC and NIPPV for AHRF (24). Specifically, NIPPV use in non-COVID-19 severe ARDS patients has been shown to result in higher ICU mortality (19), which is potentially attributable to the self-inflicted lung injury phenomenon of high spontaneous respiratory drive, generation of large tidal volumes, and heterogeneity of ventilation and regional distribution ventilation resulting in uncontrolled, potentially injurious transpulmonary pressures (25). Historically, treatment with NIPPV for H1N1 influenza and the Middle East Respiratory Syndrome-coronavirus resulted in high failure rates leading to ETI, and clear benefits of NIPPV use in SARS-CoV-1 have not been established (26). While we observed that a small subset of patients experienced good outcomes with NIPPV, further studies comparing NIPPV with other forms of respiratory support are needed. Importantly, our observation that a subset of patients using non-invasive respiratory support was successfully discharged from hospital without requiring ETI suggests that a uniform approach of early ETI for all patients with COVID-19 AHRF deserves consideration (27-30). Avoidance of unnecessary ETI and IMV is paramount to reduce IMV-associated complications, limit healthcare worker exposure associated with the ETI process (31), and preserve mechanical ventilators in resource-constrained pandemic settings. Thus, the impact of non-invasive respiratory support modalities on reducing risk for ETI and IMV and mortality merits further investigation.

Closer examination of non-invasive respiratory support use patterns revealed that the highest probabilities of requiring HFNC or NIPPV occurred relatively early in the hospital course, consistent with other studies reporting deteriorating respiratory status and development of ARDS soon after admission (4, 5, 32). Notably, we found that the median time to treatment failure was within two days after initiation of HFNC or NIPPV, highlighting that the immediate period after initiation of non-invasive respiratory support represents a crucial window for clinical deterioration and progression of disease. This is consistent with non-COVID-19 studies which have demonstrated that, for example, a ROX index (ratio of oxygen saturation/FiO_2_ to respiratory rate) (33), of ≥4.88 after 12 hours of HFNC treatment for AHRF, is associated with a significantly lower likelihood of requiring ETI (33). Furthermore, analysis of a subset of patients with non-COVID-19 ARDS treated with NIPPV from a large, multicenter observational study showed that higher severity of illness, and worse oxygenation and ventilation over the first two days of NIPPV use, were independently associated with the need for ETI (19, 34). Thus, in keeping with recommendations from the Surviving Sepsis Campaign guidelines for COVID-19 (12), close monitoring and frequent reassessment of patients receiving non-invasive respiratory support, particularly immediately after initiation, is essential.

Notably, we found that comorbid cardiovascular disease, including ischemic heart disease, congestive heart failure, and arrhythmias, was independently and robustly associated with increased risk of both HFNC and NIPPV failure. This finding is consistent with and expands upon other observational studies, which have demonstrated that cardiovascular disease is a common comorbidity among COVID-19 patients treated with non-invasive respiratory support and is associated with poor outcomes (35). This relationship has been hypothesized to stem from viral-mediated cardiac complications, including myocarditis, vascular inflammation with resultant cardiac injury and dysfunction, and new or worsening cardiac arrhythmias (4, 36-39). Our work further highlights patients with cardiovascular disease as a vulnerable subgroup that is susceptible to worse outcomes once initiated on non-invasive respiratory support.

We also observed other important clinical characteristics associated with HFNC and NIPPV outcomes. As expected, patients with higher severity of illness--as represented by more severe hypoxemia at non-invasive respiratory support initiation--were at significantly higher risk of both HFNC and NIPPV failure. Further characterization of the mechanisms and the extent to which the distribution of gas exchange and regional ventilation can be corrected by non-invasive therapies are needed to better predict disease trajectory and outcomes specific to COVID-19. Additionally, laboratory markers suggestive of more severe immune dysregulation, specifically elevated interleukin-6 and lymphopenia, were independently associated with failure in the NIPPV cohort. Interleukin-6 is a pro-inflammatory cytokine frequently upregulated in patients with severe COVID-19 as part of a dysregulated immune response, and elevated levels have been suggested to portend worse outcomes, including worsening acute respiratory failure and need for IMV (40, 41). Furthermore, viral-mediated disruption of usual immune function may cause lymphocyte exhaustion and lymphopenia (40), which has been associated with more severe disease and increased mortality (3, 5, 42). Lastly, hospital site influenced outcomes in the HFNC cohort, which may be attributable to differences in patient severity of illness, resources, or capacity strain during pandemic settings. Determination of the cause is beyond the scope of this study but deserves further exploration. Taken together, specific demographic and early clinical characteristics at the time of non-invasive respiratory support initiation may inform clinicians of susceptible subgroups of patients for whom close monitoring and early consideration of alternative management strategies is warranted if clear clinical improvement is not evident.

Our study has notable strengths. First, this is the largest longitudinal cohort study to date to evaluate outcomes in patients treated with HFNC and NIPPV on COVID-19 AHRF. Second, the granularity of our data enabled capture of accurate longitudinal data representing time-varying laboratory results, vital signs and oxygenation status, and allowed us to map outcome trajectories associated with these devices. Third, the diversity of our cohort, drawn from hospital sites representing distinct communities, improves the generalizability of our findings to other COVID-19 populations. Fourth, we explored potential bias with several sensitivity analyses, accounting for patients with missing data and the impact of anticoagulation and corticosteroids on the main analysis. Finally, we had a minimal number of censored patients, improving the reliability of our outcomes.

This study also has several limitations. As an observational study, we cannot draw conclusions on the utility of HFNC and NIPPV compared to low-flow supplemental oxygen. However, our study provides a strong basis for further efforts, including case-control studies and randomized controlled trials to investigate the efficacy of non-invasive respiratory support in reducing risks of ETI and mortality. Additionally, we chose to focus on factors at the time of HFNC or NIPPV initiation associated with treatment outcome and did not address the impact of subsequent changes in clinical characteristics or treatments administered. We were also limited to using SpO_2_/FiO_2_ ratios to approximate degree of hypoxemia (43), for which accurate interpretation of severity may be limited as arterial blood gases were unavailable for most patients. Furthermore, we did not account for HFNC flow rates and NIPPV inspiratory/expiratory pressures, which may have influenced outcomes. Finally, we did not assess healthcare worker infectious exposure risk, though prior studies of healthcare workers exposed to SARS-CoV 1 patients on HFNC or NIPPV did not report significantly higher transmission risk (44, 45) and in a COVID-19 simulation study, the exhaled air dispersion during well-fitted HFNC and CPAP use was limited (46).

## CONCLUSIONS

A subset of patients treated with HFNC and/or NIPPV achieved hospital discharge without requiring ETI and IMV, suggesting that some patients with COVID-19 AHRF can be managed effectively with these respiratory support modalities. Attention to specific demographic and early clinical factors, such as co-morbid cardiovascular disease and severity of hypoxemia, may help inform use of non-invasive respiratory strategies, allowing for a more personalized approach to the management of AHRF in pandemic settings.

## Data Availability

Deidentified data will be made publicly available.

## Acknowledgements

We would like to thank our colleague Dr. Emilia Bagiella for her statistical input on this manuscript.

## SUPPLEMENTARY APPENDIX

**Table E1.** Univariate analyses among the HFNC and NIPPV cohorts, using sub-distribution hazard ratio estimates for HFNC and NIPPV treatment failure, with live hospital discharge as a competing risk. Variable with p-value ≤0.15 were selected to be included in the multivariable model.

| Covariates | HFNC cohort<br>(N=331) |  |  | NIPPV cohort<br>(N=747) |  |  |
| --- | --- | --- | --- | --- | --- | --- |
|  | sHR (95% CI) | p-value | missing | sHR (95% CI) | p-value | missing |
| **Age** | 1.02 (1.01-1.04) | <.0001 |  | 1.01 (1.01-1.02) | 0.0001 |  |
| **Male sex** | 0.74 (0.55-1) | 0.046 |  | 0.98 (0.83-1.16) | 0.839 |  |
| **Race** |  |  |  |  |  |  |
| White | reference |  |  | reference |  |  |
| Black | 0.83 (0.58-1.19) | 0.305 |  | 0.93 (0.73-1.18) | 0.550 |  |
| Asian/Pacific Islander | 0.56 (0.24-1.28) | 0.168 |  | 0.91 (0.6-1.38) | 0.650 |  |
| Other/unknown | 0.9 (0.59-1.38) | 0.632 |  | 1.01 (0.82-1.24) | 0.920 |  |
| **Ethnicity** |  |  |  |  |  |  |
| Non-Hispanic | reference |  |  | reference |  |  |
| Hispanic | 1.7 (1.1-2.62) | 0.018 |  | 0.93 (0.78-1.12) | 0.471 |  |
| Unknown | 0.96 (0.69-1.33) | 0.811 |  | 1.16 (0.91-1.47) | 0.245 |  |
| **Insurance** |  |  |  |  |  |  |
| Medicare | reference |  |  | reference |  |  |
| Medicaid | 0.98 (0.69-1.39) | 0.892 |  | 0.81 (0.61-1.08) | 0.152 |  |
| Private | 0.63 (0.41-0.97) | 0.034 |  | 0.79 (0.62-1.01) | 0.055 |  |
| Self-pay/Other/unknown | 0.38 (0.22-0.67) | 0.001 |  | 0.72 (0.59-0.88) | 0.001 |  |
| **Co-morbidities** |  |  |  |  |  |  |
| Hypertension | 0.87 (0.65-1.17) | 0.348 |  | 1.17 (0.99-1.39) | 0.060 |  |
| Type 2 diabetes mellitus | 1.14 (0.84-1.55) | 0.410 |  | 0.89 (0.75-1.05) | 0.169 |  |
| Chronic lung disease* | 1.19 (0.87-1.62) | 0.271 |  | 0.74 (0.63-0.88) | 0.0004 |  |
| Cardiovascular disease† | 2.1 (1.54-2.88) | <.0001 |  | 1.3 (1.1-1.55) | 0.002 |  |
| Liver disease | 0.61 (0.25-1.46) | 0.268 |  | 0.94 (0.68-1.3) | 0.701 |  |
| Chronic kidney disease | 1.23 (0.85-1.79) | 0.277 |  | 1.11 (0.92-1.35) | 0.290 |  |
| Cancer | 1.25 (0.85-1.85) | 0.263 |  | 0.84 (0.65-1.09) | 0.194 |  |
| **Smoking status** |  |  |  |  |  |  |
| Never | reference |  |  | reference |  |  |
| Unknown | 0.86 (0.39-1.94) | 0.722 |  | 1.09 (0.89-1.33) | 0.412 |  |
| Former | 1 (0.7-1.43) | 0.996 |  | 1.05 (0.85-1.28) | 0.665 |  |
| Current | 0.92 (0.63-1.35) | 0.671 |  | 0.94 (0.61-1.44) | 0.771 |  |
| **Vital signs** |  |  |  |  |  |  |
| Heart rate (beats per minute) | 2.33 (1.01-5.33) | 0.046 |  | 2.3 (1.46-3.62) | 0.0003 |  |
| Systolic blood pressure | 1.48 (0.57-3.83) | 0.424 |  | 1.18 (0.72-1.92) | 0.516 |  |
| Diastolic blood pressure | 0.24 (0.1-0.59) | 0.002 |  | 0.66 (0.41-1.07) | 0.089 |  |
| Respiratory rate | 1.75 (0.94-3.26) | 0.079 |  | 1.36 (1.01-1.84) | 0.044 |  |
| SpO<sub>2</sub>/FiO<sub>2</sub> ratio | 0.41 (0.28-0.6) | <.0001 | 38 | 0.32 (0.23-0.46) | <.0001 | 52 |
| Temperature (°Fahrenheit) | 1.07 (0.97-1.19) | 0.163 |  | 1.03 (0.98-1.09) | 0.243 |  |
| **Body mass index** | 0.76 (0.38-1.55) | 0.4521 | 41 | 0.71 (0.5-1) | 0.051 | 132 |
| **Laboratory values** |  |  |  |  |  |  |
| White blood cell count (x10<sup>3</sup>/μL) | 1.06 (0.77-1.46) | 0.716 |  | 1.34 (1.11-1.62) | 0.003 |  |
| Peripheral lymphocyte (%) | 0.92 (0.72-1.18) | 0.513 | 3 | 0.73 (0.64-0.84) | <.0001 | 5 |
| Hemoglobin (g/dL) | 0.32 (0.15-0.69) | 0.004 |  | 0.66 (0.44-1) | 0.051 |  |
| Platelets (x10<sup>3</sup>/μL) | 0.61 (0.46-0.82) | 0.001 |  | 0.77 (0.64-0.92) | 0.004 |  |
| D-Dimer (μg/mL) | 1.12 (0.98-1.28) | 0.099 | 16 | 1.15 (1.07-1.25) | 0.0003 | 40 |
| C-reactive protein (mg/L) | 1.27 (1.04-1.56) | 0.021; | 13 | 1.21 (1.09-1.35) | 0.0004 | 35 |
| Ferritin (ng/mL) | 1.12 (0.98-1.28) | 0.103 | 11 | 1.07 (0.99-1.15) | 0.070 | 37 |
| Interleukin-6 (pg/mL) | 1.37 (1.18-1.59) | <.0001 | 72 | 1.27 (1.17-1.39) | <.0001 | 232 |
| Procalcitonin (ng/mL) | 1.15 (1.04-1.26) | 0.005 | 21 | 1.1 (1.04-1.16) | 0.0004 | 82 |
| Creatinine (mg/dL) | 1.17 (0.97-1.41) | 0.107 |  | 1.15 (1.03-1.29) | 0.014 |  |
| **Code status<sup>†</sup>** |  |  |  |  |  |  |
| Do-not-intubate, Yes | 1.75 (1.27-2.4) | 0.001 |  | 1.2 (0.98-1.48) | 0.082 |  |
| **Treatments received<sup>§</sup>** |  |  |  |  |  |  |
| Prophylactic anticoagulation, Yes | 1.57 (1.12-2.21) | 0.010 |  | 0.96 (0.81-1.13) | 0.596 |  |
| Therapeutic anticoagulation, Yes | 0.88 (0.63-1.21) | 0.424 |  | 1.01 (0.83-1.22) | 0.931 |  |
| Corticosteroids, Yes | 1.29 (0.4-4.18) | 0.669 |  | 1.23 (0.81-1.86) | 0.330 |  |
| **Hospital admission site** |  |  |  |  |  |  |
| Quaternary referral hospital | reference |  |  | reference |  |  |
| Community hospital A | 0.27 (0.14-0.5) | <.0001 |  | 1.44 (1.14-1.8) | 0.002 |  |
| Community hospital B | 0.63 (0.43-0.94) | 0.024 |  | 1.58 (1.19-2.08) | 0.001 |  |
| Community hospital C | 0.94 (0.57-1.57) | 0.816 |  | 0.96 (0.76-1.22) | 0.736 |  |
| Tertiary hospital | 0.82 (0.51-1.32) | 0.409 |  | 0.97 (0.72-1.29) | 0.821 |  |

\*Chronic lung disease includes: Asthma, emphysema, COPD, chronic bronchitis, bronchiectasis, atelectasis, diaphragmatic disease, interstitial lung diseases, sarcoidosis, and disorders of mediastinum.
<sup>†</sup>Cardiovascular disease includes: Atherosclerotic heart disease, ischemic heart disease, congestive heart failure, rheumatic heart disease, pericardial disease, myocarditis, endocarditis, valvular disorders, cardiomyopathy, arrhythmias, history of cardiac arrest, peripheral vascular disease, aortic aneurysm, orthostatic hypotension, pulmonary hypertension, cardiac arrest, post-procedural cardiac complications.
<sup>‡</sup>At time of initiation of HFNC or NIPPV.
<sup>§</sup>From hospital admission to initiation of HFNC or NIPPV.

**Table E2.**
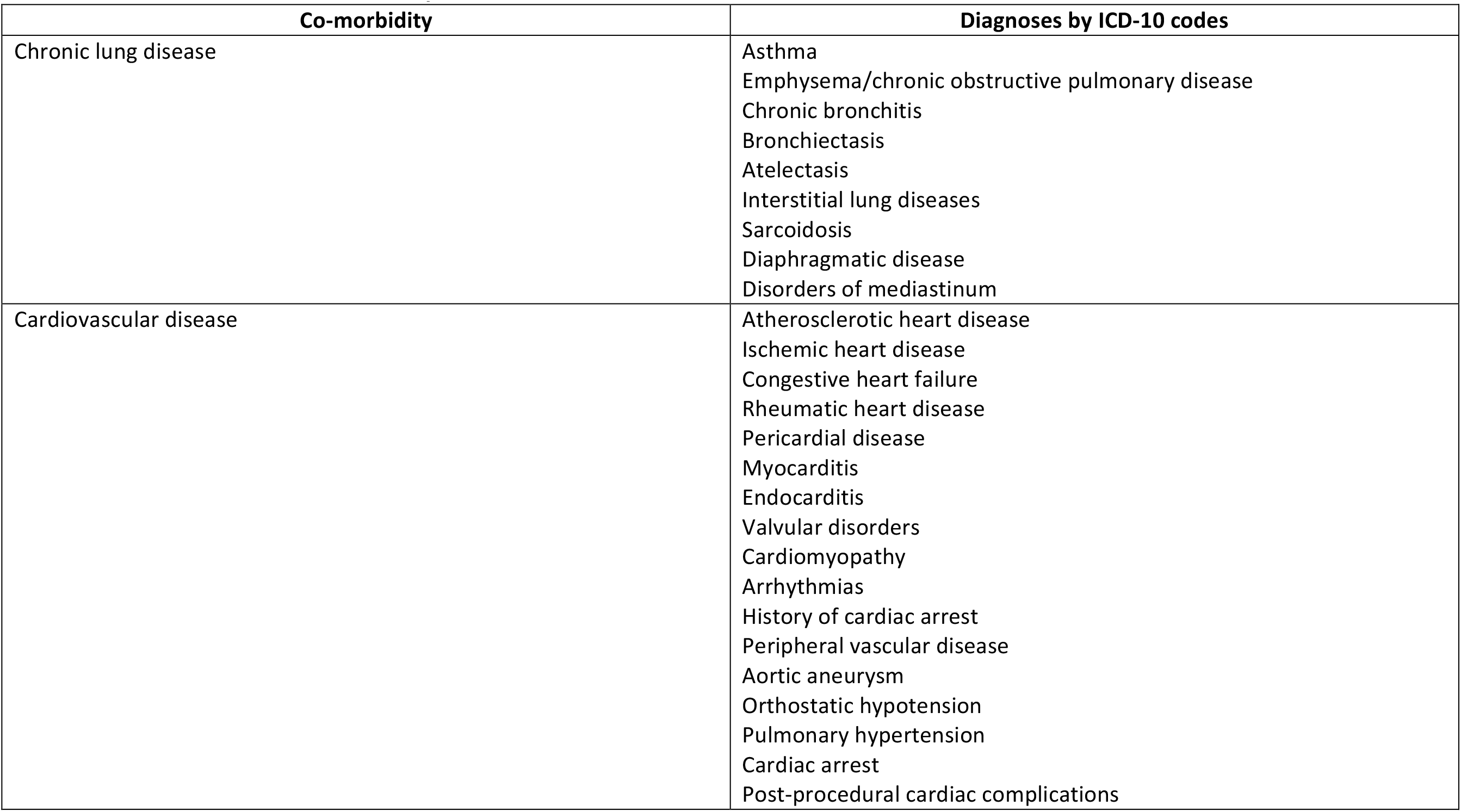
Breakdown of co-morbidities by ICD-10 codes.

**Table E3.** Sensitivity analysis results among the HFNC cohort from three models: 1) adding receipt of prophylactic-dose and therapeutic-dose anticoagulation at HFNC initiation as a covariate; 2) adding receipt of corticosteroids at HFNC initiation as a covariate; and 3) removing IL-6 and BMI resulting in a larger subset of patients, using sub-distribution hazard ratio estimates for HFNC treatment failure, with live hospital discharge as a competing risk. Highlighted rows are covariates, which remained significant throughout the main and all sensitivity multivariable models.

| Covariates | Model 1 (n=232)<br>sHR (95% CI) | p-value | Model 2 (n=232)<br>sHR (95% CI) | p-value | Model 3 (n=271)<br>sHR (95% CI) | p-value |
| --- | --- | --- | --- | --- | --- | --- |
| Age (years) | 1 (0.99 - 1.02) | 0.841 | 1 (0.99 - 1.02) | 0.747 | 1.01 (0.99 - 1.02) | 0.283 |
| Male sex (vs. Female) | 0.97 (0.65 - 1.46) | 0.891 | 0.87 (0.59 - 1.28) | 0.475 | 0.89 (0.62 - 1.28) | 0.528 |
| Ethnicity |  |  |  |  |  |  |
| Non-Hispanic | Reference |  | Reference |  | Reference |  |
| Hispanic | 1 (0.63 - 1.58) | 0.997 | 1.07 (0.68 - 1.69) | 0.759 | 0.83 (0.55 - 1.24) | 0.354 |
| Unknown | 1.45 (0.81 - 2.62) | 0.212 | 1.68 (0.98 - 2.88) | 0.061 | 1.68 (1.04 - 2.72) | 0.035 |
| Insurance |  |  |  |  |  |  |
| Medicare | Reference |  | Reference |  | Reference |  |
| Medicaid | 0.53 (0.24 - 1.16) | 0.114 | 0.46 (0.2 - 1.03) | 0.060 | 0.54 (0.25 - 1.19) | 0.126 |
| Private | 0.85 (0.47 - 1.56) | 0.603 | 0.79 (0.43 - 1.45) | 0.441 | 0.81 (0.46 - 1.4) | 0.443 |
| Self-pay/Other/unknown | 1.04 (0.61 - 1.77) | 0.884 | 0.91 (0.53 - 1.56) | 0.722 | 1.01 (0.66 - 1.55) | 0.966 |
| Comorbidities |  |  |  |  |  |  |
| Cardiovascular disease* (Yes vs. No) | 1.96 (1.25 - 3.08) | 0.003 | 1.87 (1.16 - 3) | 0.010 | 1.75 (1.15 - 2.66) | 0.009 |
| Cerebrovascular disease (Yes vs. No) | 1.7 (1 - 2.9) | 0.050 | 1.34 (0.76 - 2.36) | 0.309 | 1.66 (1.01 - 2.72) | 0.045 |
| Vital signs |  |  |  |  |  |  |
| Heart rate (beats per minute) | 1.85 (0.52 - 6.6) | 0.343 | 1.97 (0.55 - 7.08) | 0.301 | 2.58 (1 - 6.68) | 0.050 |
| Systolic blood pressure |  |  |  |  |  |  |
| Diastolic blood pressure | 0.28 (0.08 - 0.92) | 0.036 | 0.32 (0.09 - 1.1) | 0.071 | 0.3 (0.1 - 0.96) | 0.042 |
| Respiratory rate | 2.1 (0.82 - 5.35) | 0.121 | 1.93 (0.78 - 4.78) | 0.156 | 1.35 (0.57 - 3.18) | 0.493 |
| SpO<sub>2</sub>/FiO<sub>2</sub> ratio | 0.35 (0.21 - 0.6) | 0.0001 | 0.31 (0.18 - 0.54) | <.0001 | 0.3 (0.18 - 0.52) | <.0001 |
| Laboratory values |  |  |  |  |  |  |
| Hemoglobin (g/dL) | 0.63 (0.19 - 2.11) | 0.454 | 0.53 (0.16 - 1.73) | 0.294 | 0.49 (0.17 - 1.46) | 0.202 |
| Platelets (x10<sup>3</sup>/μL) | 0.71 (0.48 - 1.04) | 0.079 | 0.63 (0.43 - 0.91) | 0.014 | 0.65 (0.46 - 0.92) | 0.014 |
| D-Dimer (μg/mL) | 0.93 (0.76 - 1.14) | 0.506 | 0.9 (0.75 - 1.09) | 0.296 | 0.87 (0.73 - 1.04) | 0.122 |
| C-reactive protein (mg/L) | 1.14 (0.86 - 1.52) | 0.356 | 1.21 (0.91 - 1.61) | 0.190 | 1.4 (1.08 - 1.81) | 0.010 |
| Ferritin (ng/mL) | 0.9 (0.73 - 1.12) | 0.347 | 0.93 (0.74 - 1.16) | 0.510 | 0.99 (0.82 - 1.18) | 0.864 |
| Interleukin-6 (pg/mL) | 1.15 (0.96 - 1.38) | 0.131 | 1.15 (0.96 - 1.37) | 0.134 | - | - |
| Pro-calcitonin (ng/mL) | 1.01 (0.86 - 1.18) | 0.935 | 1.01 (0.86 - 1.2) | 0.884 | 1 (0.86 - 1.16) | 0.981 |
| Creatinine (mg/dL) | 1 (0.71 - 1.41) | 0.983 | 0.93 (0.66 - 1.31) | 0.679 | 0.93 (0.68 - 1.27) | 0.649 |
| Code status<sup>†</sup> |  |  |  |  |  |  |
| Do-not-intubate (Yes vs. No) | 0.77 (0.48 - 1.25) | 0.296 | 0.78 (0.48 - 1.28) | 0.328 | 0.77 (0.49 - 1.21) | 0.251 |

| Treatments received<sup>†</sup> |  |  |  |  |  |  |
| --- | --- | --- | --- | --- | --- | --- |
| Prophylactic anticoagulation (Yes vs. No) | 2.05 (1.25 - 3.37) | 0.005 | - | - | - | - |
| Therapeutic anticoagulation (Yes vs. No) | 0.78 (0.49 - 1.23) | 0.283 | - | - | - | - |
| Corticosteroids (Yes vs. No) | - | - | 1.57 (0.37 - 6.63) | 0.544 | - | - |
| Hospital admission site, no. (%) |  |  |  |  |  |  |
| Quaternary referral hospital | Reference |  | Reference |  | Reference |  |
| Community hospital A | 1.18 (0.52 - 2.67) | 0.700 | 1.25 (0.57 - 2.71) | 0.578 | 1.22 (0.66 - 2.23) | 0.524 |
| Community hospital B | 1.35 (0.31 - 5.87) | 0.693 | 1.07 (0.21 - 5.36) | 0.939 | 1.15 (0.52 - 2.54) | 0.735 |
| Community hospital C | 0.51 (0.28 - 0.92) | 0.025 | 0.56 (0.31 - 1) | 0.048 | 0.46 (0.26 - 0.81) | 0.007 |
| Tertiary hospital | 0.23 (0.12 - 0.46) | <.0001 | 0.3 (0.16 - 0.56) | 0.000 | 0.26 (0.14 - 0.48) | <.0001 |

\* Cardiovascular disease includes: Atherosclerotic heart disease, ischemic heart disease, congestive heart failure, rheumatic heart disease, pericardial disease, myocarditis, endocarditis, valvular disorders, cardiomyopathy, arrhythmias, history of cardiac arrest, peripheral vascular disease, aortic aneurysm, orthostatic hypotension, pulmonary hypertension, cardiac arrest, post-procedural cardiac complications.
<sup>†</sup>At time of initiation of HFNC or NIPPV.
<sup>‡</sup>From hospital admission to initiation of HFNC or NIPPV.
sHR: Sub-distribution hazard ratio; CI: Confidence interval; HFNC: High-flow oxygen through nasal cannula; SpO<sub>2</sub>: Oxygen saturation; FiO<sub>2</sub>: Fraction of inspired oxygen. All continuous variables, except for age, were natural log-transformed.

**Table E4.** Sensitivity analysis results among the NIPPV cohort from three models: 1) adding receipt of prophylactic-dose and therapeutic-dose anticoagulation at NIPPV initiation as a covariate; 2) adding receipt of corticosteroids at NIPPV initiation as a covariate; and 3) removing IL-6 and BMI as covariates resulting in a larger subset of patients, using sub-distribution hazard ratio estimates for treatment failure, with live hospital discharge as a competing risk. Highlighted rows are covariates, which remained significant throughout the main and all sensitivity multivariable models.

| Covariates | Model 1 (n=408)<br>sHR (95% CI) | p-value | Model 2 (n=408)<br>sHR (95% CI) | p-value | Model 3 (n=605)<br>sHR (95% CI) | p-value |
| --- | --- | --- | --- | --- | --- | --- |
| Age (years) | 1 (0.99 - 1.02) | 0.5619 | 1 (0.99 - 1.01) | 0.7079 | 1.01 (1 - 1.02) | 0.0811 |
| Insurance |  |  |  |  |  |  |
| Medicare | Reference |  | Reference |  | Reference |  |
| Medicaid | 0.7 (0.43 - 1.15) | 0.1632 | 0.71 (0.43 - 1.17) | 0.1753 | 0.79 (0.55 - 1.15) | 0.2158 |
| Private | 0.94 (0.63 - 1.41) | 0.7689 | 0.92 (0.61 - 1.37) | 0.6648 | 0.92 (0.68 - 1.27) | 0.6239 |
| Self-pay/Other/unknown | 0.83 (0.61 - 1.13) | 0.2436 | 0.8 (0.59 - 1.09) | 0.1628 | 0.73 (0.57 - 0.94) | 0.0134 |
| Comorbidities |  |  |  |  |  |  |
| Hypertension (Yes vs. No) | 1.08 (0.82 - 1.42) | 0.6059 | 1.07 (0.81 - 1.42) | 0.6119 | 1.12 (0.91 - 1.39) | 0.2964 |
| Chronic lung disease* (Yes vs. No) | 0.9 (0.68 - 1.19) | 0.4546 | 0.89 (0.67 - 1.16) | 0.3813 | 0.84 (0.68 - 1.03) | 0.0866 |
| Cardiovascular disease<sup>†</sup> (Yes vs. No) | 1.41 (1.07 - 1.85) | 0.0136 | 1.4 (1.06 - 1.84) | 0.0162 | 1.23 (1 - 1.51) | 0.0493 |
| Vital signs |  |  |  |  |  |  |
| Heart rate (beats per minute) | 1.23 (0.52 - 2.92) | 0.6385 | 1.24 (0.52 - 2.93) | 0.6318 | 1.87 (1.03 - 3.38) | 0.0393 |
| Diastolic blood pressure | 0.49 (0.23 - 1.05) | 0.0663 | 0.5 (0.23 - 1.07) | 0.0731 | 0.64 (0.38 - 1.09) | 0.0996 |
| Respiratory rate | 1.19 (0.8 - 1.78) | 0.3897 | 1.17 (0.79 - 1.74) | 0.4386 | 1.25 (0.89 - 1.77) | 0.202 |
| SpO2/FiO2 ratio | 0.34 (0.21 - 0.54) | <.0001 | 0.34 (0.21 - 0.55) | <.0001 | 0.33 (0.22 - 0.49) | <.0001 |
| Body mass index | 0.87 (0.53 - 1.42) | 0.5746 | 0.83 (0.51 - 1.36) | 0.4657 | - | - |
| Laboratory values |  |  |  |  |  |  |
| White blood cell count (x10<sup>3</sup>/μL) | 1.06 (0.76 - 1.47) | 0.7411 | 1.07 (0.76 - 1.49) | 0.7056 | 1.08 (0.84 - 1.39) | 0.5456 |
| Peripheral lymphocyte (%) | 0.73 (0.58 - 0.92) | 0.0082 | 0.74 (0.59 - 0.94) | 0.0125 | 0.82 (0.68 - 0.99) | 0.0358 |
| Hemoglobin (g/dL) | 0.39 (0.18 - 0.85) | 0.018 | 0.41 (0.19 - 0.88) | 0.0218 | 0.67 (0.39 - 1.14) | 0.14 |
| Platelets (x10<sup>3</sup>/μL) | 0.77 (0.58 - 1.02) | 0.0652 | 0.77 (0.58 - 1.02) | 0.0659 | 0.71 (0.58 - 0.88) | 0.0017 |
| D-Dimer (μg/mL) | 1.01 (0.89 - 1.15) | 0.8847 | 1.01 (0.88 - 1.15) | 0.938 | 1.02 (0.93 - 1.13) | 0.6662 |
| C-reactive protein (mg/L) | 1.1 (0.94 - 1.28) | 0.2525 | 1.11 (0.95 - 1.29) | 0.1998 | 1.13 (1.01 - 1.27) | 0.0348 |
| Ferritin (ng/mL) | 1.08 (0.96 - 1.23) | 0.2089 | 1.07 (0.94 - 1.21) | 0.2921 | 1.03 (0.93 - 1.13) | 0.6268 |
| Interleukin-6 (pg/mL) | 1.13 (1.02 - 1.26) | 0.0252 | 1.13 (1.02 - 1.26) | 0.0212 | - | - |
| Pro-calcitonin (ng/mL) | 0.96 (0.87 - 1.07) | 0.4965 | 0.97 (0.87 - 1.07) | 0.5062 | 1 (0.93 - 1.08) | 0.9075 |
| Creatinine (mg/dL) | 0.9 (0.72 - 1.12) | 0.3298 | 0.91 (0.73 - 1.13) | 0.4017 | 0.98 (0.84 - 1.14) | 0.754 |
| **Code status<sup>†</sup>** |  |  |  |  |  |  |
| Do-not-intubate (Yes vs. No) | 0.91 (0.65 - 1.27) | 0.5641 | 0.88 (0.63 - 1.24) | 0.4639 | 1.03 (0.8 - 1.33) | 0.8016 |
| **Treatments received<sup>§</sup>** |  |  |  |  |  |  |
| Prophylactic anticoagulation (Yes vs. No) | 1.1 (0.85 - 1.42) | 0.4783 | - | - | - | - |
| Therapeutic anticoagulation (Yes vs. No) | 0.84 (0.62 - 1.12) | 0.2264 | - | - | - | - |
| Corticosteroids (Yes vs. No) | - | - | 0.96 (0.52 - 1.76) | 0.8893 | - | - |
| **Hospital admission site** |  |  |  |  |  |  |
| Quaternary referral hospital | Reference |  | Reference |  | Reference |  |
| Community hospital A | 1.08 (0.71 - 1.65) | 0.7241 | 1.09 (0.72 - 1.65) | 0.68 | 1.24 (0.94 - 1.63) | 0.1279 |
| Community hospital B | 1.59 (0.89 - 2.85) | 0.119 | 1.67 (0.95 - 2.94) | 0.0775 | 1.45 (1 - 2.09) | 0.0496 |
| Community hospital C | 0.97 (0.69 - 1.35) | 0.8483 | 0.99 (0.71 - 1.39) | 0.9687 | 0.91 (0.69 - 1.19) | 0.4917 |
| Tertiary hospital | 0.75 (0.52 - 1.1) | 0.1372 | 0.8 (0.56 - 1.15) | 0.2294 | 0.82 (0.6 - 1.14) | 0.2452 |

\*Chronic lung disease includes: Asthma, emphysema, COPD, chronic bronchitis, bronchiectasis, atelectasis, diaphragmatic disease, interstitial lung diseases, sarcoidosis, and disorders of mediastinum.
<sup>†</sup>Cardiovascular disease includes: Atherosclerotic heart disease, ischemic heart disease, congestive heart failure, rheumatic heart disease, pericardial disease, myocarditis, endocarditis, valvular disorders, cardiomyopathy, arrhythmias, history of cardiac arrest, peripheral vascular disease, aortic aneurysm, orthostatic hypotension, pulmonary hypertension, cardiac arrest, post-procedural cardiac complications.
<sup>‡</sup>At time of initiation of HFNC or NIPPV.
<sup>§</sup>From hospital admission to initiation of HFNC or NIPPV.
sHR: Sub-distribution hazard ratio; CI: Confidence interval; NIPPV: Non-invasive positive pressure ventilation; SpO<sub>2</sub>: Oxygen saturation; FiO<sub>2</sub>: Fraction of inspired oxygen. All continuous variables, except for age, were natural log-transformed.

**Table E5.**
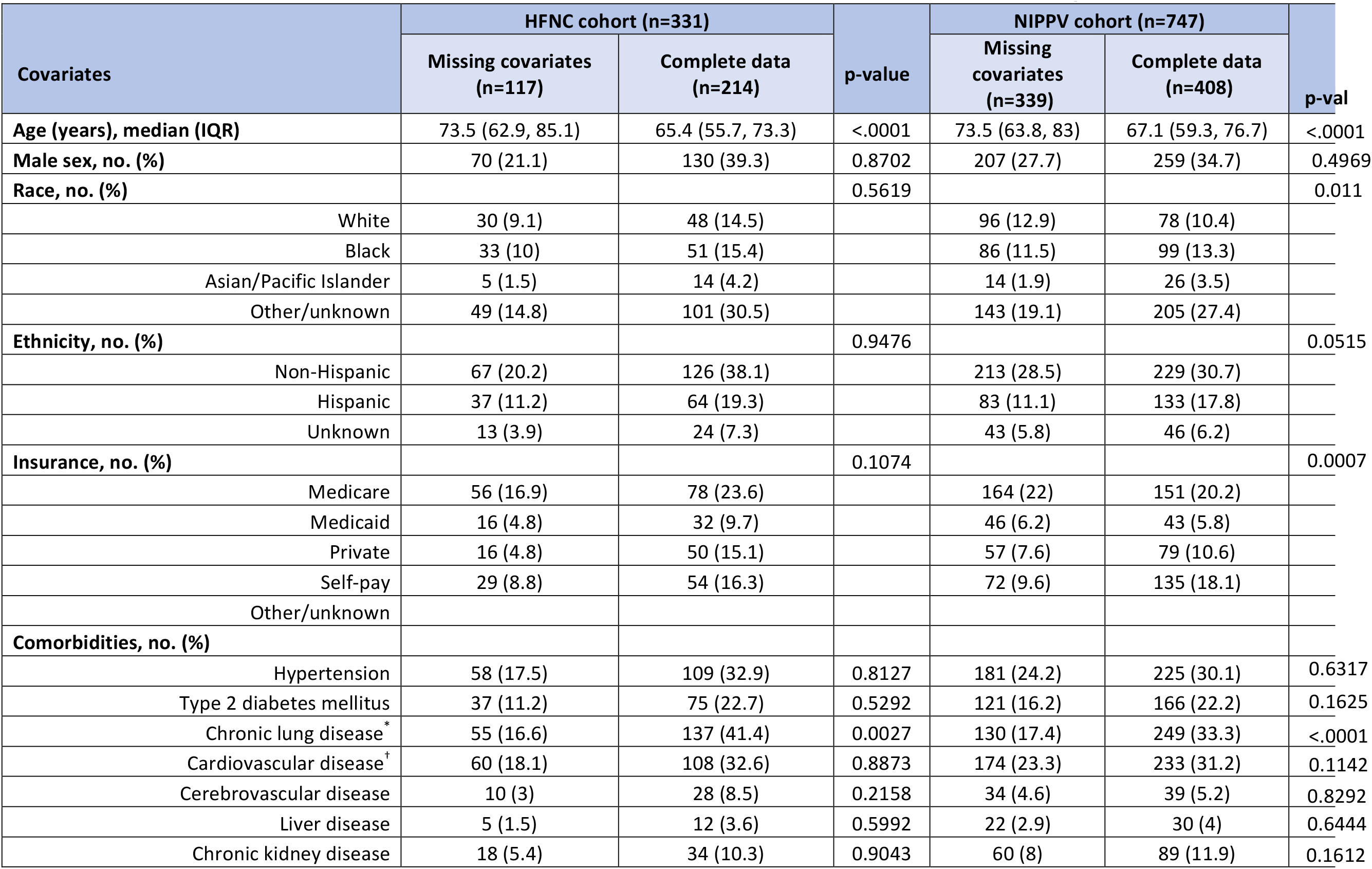
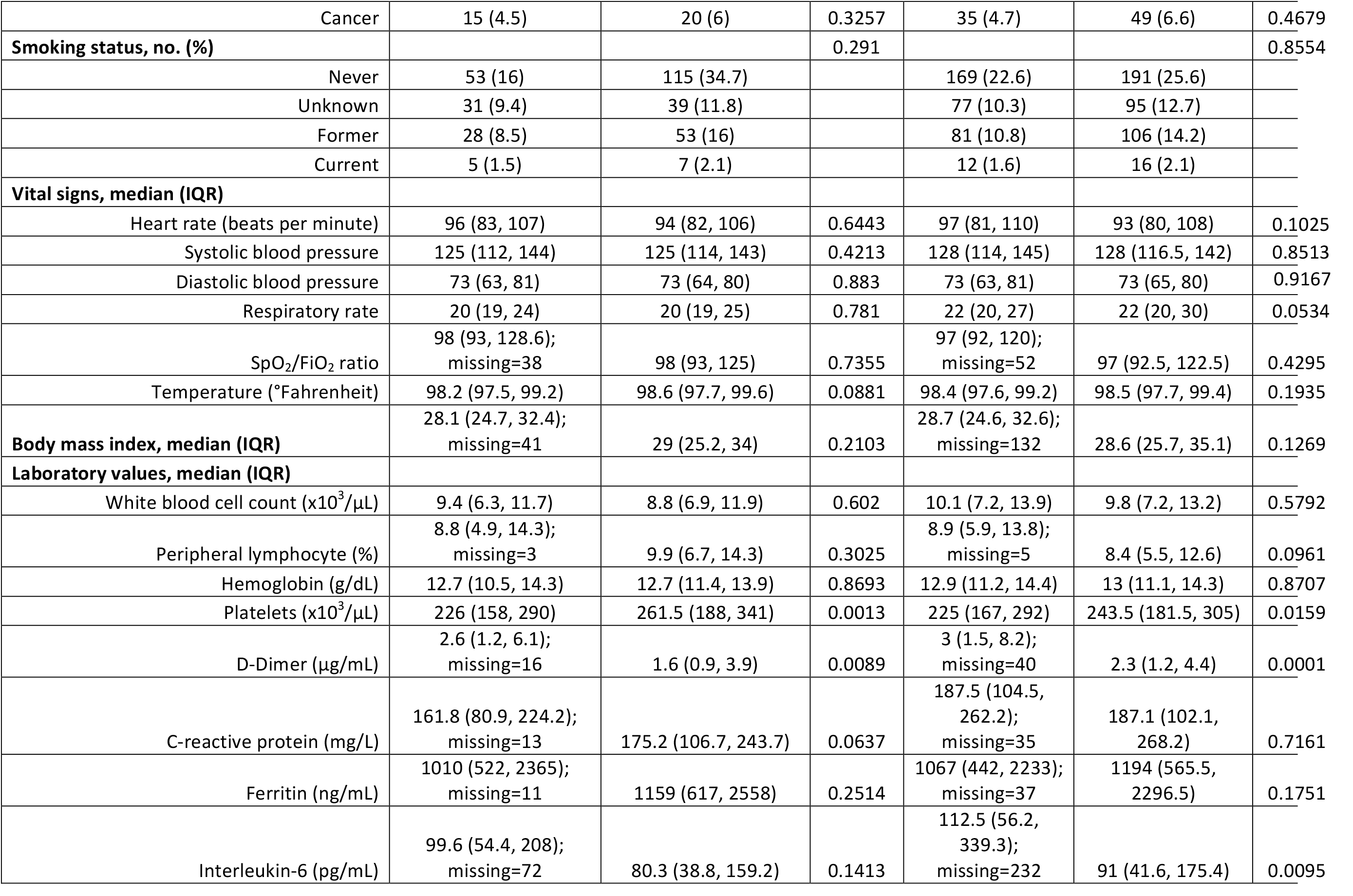
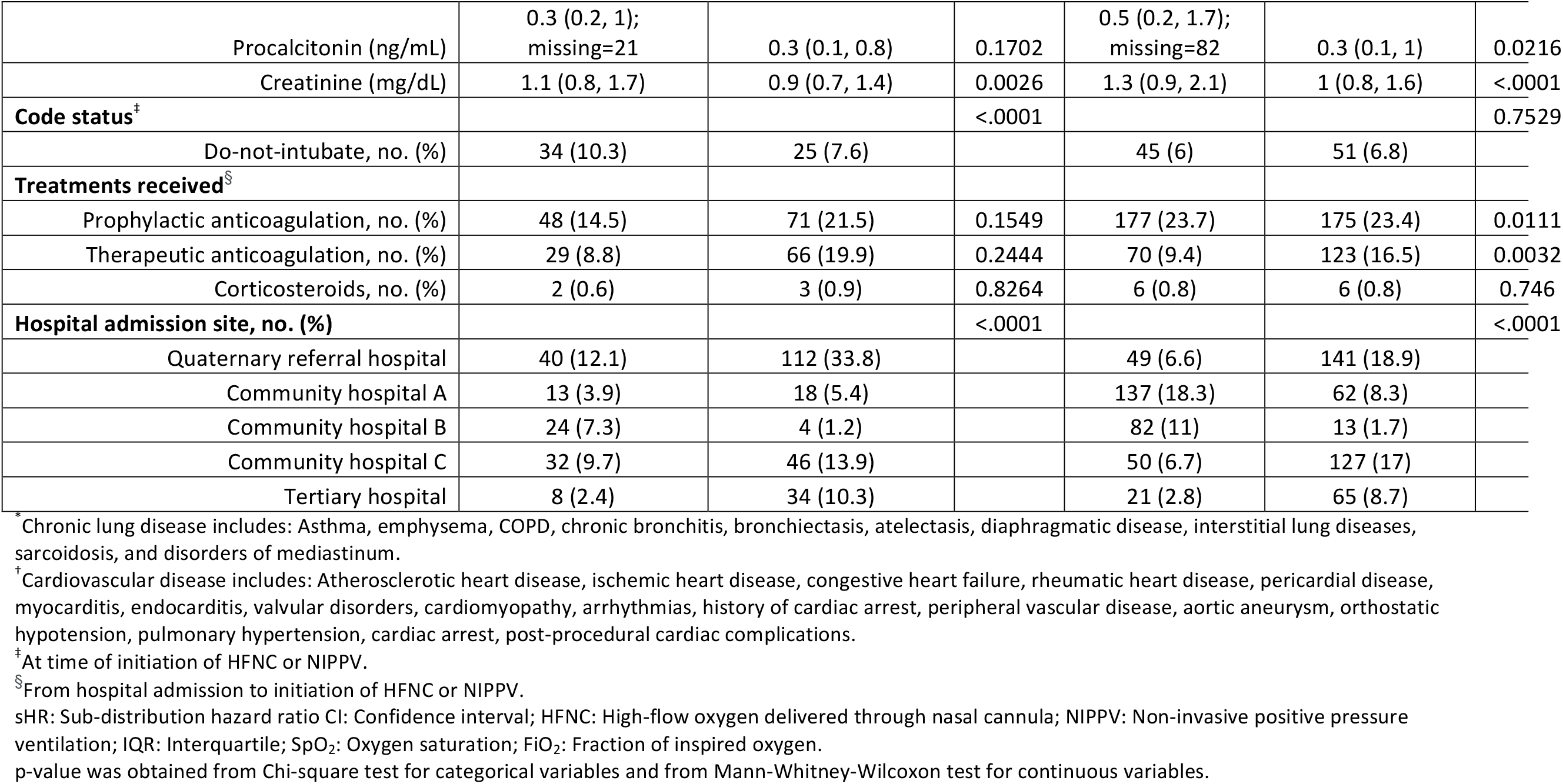
Comparisons of patient demographic and clinical characteristics between those with, versus without, any missing covariates.

## REFERENCES

1. World Health Organization. Coronavirus disease (COVID-19) pandemic. https://www.who.int/emergencies/diseases/novel-coronavirus-2019. July 2, 2020].

2. Centers for Disease Control and Prevention. CDC COVID Data Tracker. https://www.cdc.gov/covid-data-tracker/#cases. July 2, 2020].

3. Wu C, Chen X, Cai Y, Xia J, Zhou X, Xu S, et al. Risk Factors Associated With Acute Respiratory Distress Syndrome and Death in Patients With Coronavirus Disease 2019 Pneumonia in Wuhan, China. JAMA Intern Med 2020.

4. Zhou F, Yu T, Du R, Fan G, Liu Y, Liu Z, et al. Clinical course and risk factors for mortality of adult inpatients with COVID-19 in Wuhan, China: a retrospective cohort study. The Lancet 2020; 395: 1054–1062.

5. Wang D, Hu B, Hu C, Zhu F, Liu X, Zhang J, et al. Clinical Characteristics of 138 Hospitalized Patients With 2019 Novel Coronavirus-Infected Pneumonia in Wuhan, China. JAMA 2020.

6. Huang C, Wang Y, Li X, Ren L, Zhao J, Hu Y, et al. Clinical features of patients infected with 2019 novel coronavirus in Wuhan, China. The Lancet 2020; 395: 497–506.

7. Richardson S, Hirsch JS, Narasimhan M, Crawford JM, McGinn T, Davidson KW, et al. Presenting Characteristics, Comorbidities, and Outcomes Among 5700 Patients Hospitalized With COVID-19 in the New York City Area. JAMA 2020.

8. Goyal P, Choi JJ, Pinheiro LC, Schenck EJ, Chen R, Jabri A, et al. Clinical Characteristics of Covid-19 in New York City. N Engl J Med 2020.

9. Cummings MJ, Baldwin MR, Abrams D, Jacobson SD, Meyer BJ, Balough EM, et al. Epidemiology, clinical course, and outcomes of critically ill adults with COVID-19 in New York City: a prospective cohort study. The Lancet 2020.

10. Yang X, Yu Y, Xu J, Shu H, Xia Ja, Liu H, et al. Clinical course and outcomes of critically ill patients with SARS-CoV-2 pneumonia in Wuhan, China: a single-centered, retrospective, observational study. The Lancet Respiratory Medicine 2020; 8: 475–481.

11. Brewster DJ, Chrimes N, Do TB, Fraser K, Groombridge CJ, Higgs A, et al. Consensus statement: Safe Airway Society principles of airway management and tracheal intubation specific to the COVID-19 adult patient group. Med J Aust 2020.

12. Alhazzani W, Moller MH, Arabi YM, Loeb M, Gong MN, Fan E, et al. Surviving Sepsis Campaign: Guidelines on the Management of Critically Ill Adults with Coronavirus Disease 2019 (COVID-19). Crit Care Med 2020; 48: e440–e469.

13. Mauri T, Turrini C, Eronia N, Grasselli G, Volta CA, Bellani G, et al. Physiologic Effects of High-Flow Nasal Cannula in Acute Hypoxemic Respiratory Failure. American Journal of Respiratory and Critical Care Medicine 2017; 195: 1207–1215.

14. Rochwerg B, Granton D, Wang DX, Helviz Y, Einav S, Frat JP, et al. High flow nasal cannula compared with conventional oxygen therapy for acute hypoxemic respiratory failure: a systematic review and meta-analysis. Intensive Care Med 2019; 45: 563– 572.

15. Frat JP, Thille AW, Mercat A, Girault C, Ragot S, Perbet S, et al. High-flow oxygen through nasal cannula in acute hypoxemic respiratory failure. N Engl J Med 2015; 372: 2185–2196.

16. Ferreyro BL, Angriman F, Munshi L, Del Sorbo L, Ferguson ND, Rochwerg B, et al. Association of Noninvasive Oxygenation Strategies With All-Cause Mortality in Adults With Acute Hypoxemic Respiratory Failure: A Systematic Review and Meta- analysis. JAMA 2020.

17. Kang BJ, Koh Y, Lim CM, Huh JW, Baek S, Han M, et al. Failure of high-flow nasal cannula therapy may delay intubation and increase mortality. Intensive Care Med 2015; 41: 623–632.

18. Demoule A, Girou E, Richard JC, Taille S, Brochard L. Benefits and risks of success or failure of noninvasive ventilation. Intensive Care Med 2006; 32: 1756–1765.

19. Bellani G, Laffey JG, Pham T, Madotto F, Fan E, Brochard L, et al. Noninvasive Ventilation of Patients with Acute Respiratory Distress Syndrome. Insights from the LUNG SAFE Study. American Journal of Respiratory and Critical Care Medicine 2017; 195: 67–77.

20. Wang K, Zhao W, Li J, Shu W, Duan J. The experience of high-flow nasal cannula in hospitalized patients with 2019 novel coronavirus-infected pneumonia in two hospitals of Chongqing, China. Ann Intensive Care 2020; 10: 37.

21. Geng S, Mei Q, Zhu C, Yang T, Yang Y, Fang X, et al. High flow nasal cannula is a good treatment option for COVID-19. Heart Lung 2020.

22. Grasselli G, Zangrillo A, Zanella A, Antonelli M, Cabrini L, Castelli A, et al. Baseline Characteristics and Outcomes of 1591 Patients Infected With SARS-CoV-2 Admitted to ICUs of the Lombardy Region, Italy. JAMA 2020.

23. Schünemann HJ, Khabsa J, Solo K, Khamis AM, Brignardello-Petersen R, El-Harakeh A, et al. Ventilation Techniques and Risk for Transmission of Coronavirus Disease, Including COVID-19. Annals of Internal Medicine 2020.

24. Drake MG. High-Flow Nasal Cannula Oxygen in Adults: An Evidence-based Assessment. Annals of the American Thoracic Society 2018; 15: 145–155.

25. Brochard L, Slutsky A, Pesenti A. Mechanical Ventilation to Minimize Progression of Lung Injury in Acute Respiratory Failure. American Journal of Respiratory and Critical Care Medicine 2017; 195: 438–442.

26. Esquinas AM, Egbert Pravinkumar S, Scala R, Gay P, Soroksky A, Girault C, et al. Noninvasive mechanical ventilation in high-risk pulmonary infections: a clinical review. European Respiratory Review 2014; 23: 427–438.

27. Bos LD. COVID-19 Related Acute Respiratory Distress Syndrome: Not so Atypical. American Journal of Respiratory and Critical Care Medicine 2020; 0: null.

28. Gattinoni L, Chiumello D, Caironi P, Busana M, Romitti F, Brazzi L, et al. COVID-19 pneumonia: different respiratory treatments for different phenotypes? Intensive Care Med 2020.

29. Gattinoni L, Coppola S, Cressoni M, Busana M, Rossi S, Chiumello D. COVID-19 Does Not Lead to a “Typical” Acute Respiratory Distress Syndrome. American Journal of Respiratory and Critical Care Medicine 2020; 201: 1299–1300.

30. Camporota L, Vasques F, Sanderson B, Barrett NA, Gattinoni L. Identification of pathophysiological patterns for triage and respiratory support in COVID-19. The Lancet Respiratory Medicine 2020.

31. Weissman DN, de Perio MA, Radonovich LJ, Jr. COVID-19 and Risks Posed to Personnel During Endotracheal Intubation. JAMA 2020; 323: 2027–2028.

32. Wang F, Qu M, Zhou X, Zhao K, Lai C, Tang Q, et al. The timeline and risk factors of clinical progression of COVID-19 in Shenzhen, China. Journal of Translational Medicine 2020; 18: 270.

33. Roca O, Caralt B, Messika J, Samper M, Sztrymf B, Hernández G, et al. An Index Combining Respiratory Rate and Oxygenation to Predict Outcome of Nasal High-Flow Therapy. American Journal of Respiratory and Critical Care Medicine 2019; 199: 1368– 1376.

34. Bellani G, Laffey JG, Pham T, Fan E, Brochard L, Esteban A, et al. Epidemiology, Patterns of Care, and Mortality for Patients With Acute Respiratory Distress Syndrome in Intensive Care Units in 50 Countries. JAMA 2016; 315: 788–800.

35. Nishiga M, Wang DW, Han Y, Lewis DB, Wu JC. COVID-19 and cardiovascular disease: from basic mechanisms to clinical perspectives. Nature Reviews Cardiology 2020.

36. Inciardi RM, Adamo M, Lupi L, Cani DS, Di Pasquale M, Tomasoni D, et al. Characteristics and outcomes of patients hospitalized for COVID-19 and cardiac disease in Northern Italy. European Heart Journal 2020; 41: 1821–1829.

37. Mehra MR, Desai SS, Kuy S, Henry TD, Patel AN. Cardiovascular Disease, Drug Therapy, and Mortality in Covid-19. New England Journal of Medicine 2020; 382: e102.

38. Guo T, Fan Y, Chen M, Wu X, Zhang L, He T, et al. Cardiovascular Implications of Fatal Outcomes of Patients With Coronavirus Disease 2019 (COVID-19). JAMA Cardiology 2020; 5: 811–818.

39. Madjid M, Safavi-Naeini P, Solomon SD, Vardeny O. Potential Effects of Coronaviruses on the Cardiovascular System: A Review. JAMA Cardiology 2020; 5: 831–840.

40. Yang L, Liu S, Liu J, Zhang Z, Wan X, Huang B, et al. COVID-19: immunopathogenesis and Immunotherapeutics. Signal Transduction and Targeted Therapy 2020; 5: 128.

41. Herold T, Jurinovic V, Arnreich C, Lipworth BJ, Hellmuth JC, von Bergwelt-Baildon M, et al. Elevated levels of IL-6 and CRP predict the need for mechanical ventilation in COVID-19. J Allergy Clin Immunol 2020; 146: 128–136 e124.

42. Guan WJ, Ni ZY, Hu Y, Liang WH, Ou CQ, He JX, et al. Clinical Characteristics of Coronavirus Disease 2019 in China. N Engl J Med 2020; 382: 1708–1720.

43. Rice TW, Wheeler AP, Bernard GR, Hayden DL, Schoenfeld DA, Ware LB, et al. Comparison of the SpO2/FIO2 ratio and the PaO2/FIO2 ratio in patients with acute lung injury or ARDS. Chest 2007; 132: 410–417.

44. Raboud J, Shigayeva A, McGeer A, Bontovics E, Chapman M, Gravel D, et al. Risk factors for SARS transmission from patients requiring intubation: a multicentre investigation in Toronto, Canada. PLoS One 2010; 5: e10717.

45. Fowler RA, Guest CB, Lapinsky SE, Sibbald WJ, Louie M, Tang P, et al. Transmission of severe acute respiratory syndrome during intubation and mechanical ventilation. Am J Respir Crit Care Med 2004; 169: 1198–1202.

46. Hui DS, Chow BK, Ng SS, Chu LCY, Hall SD, Gin T, et al. Exhaled air dispersion distances during noninvasive ventilation via different Respironics face masks. Chest 2009; 136: 998–1005.

